# Cardiac Arrhythmia after COVID-19 Vaccination versus Non–COVID-19 Vaccination: A Systematic Review and Meta-Analysis

**DOI:** 10.1101/2022.11.21.22282554

**Authors:** Ao Shi, Xiaoyi Tang, Panpan Xia, Meiqi Hao, Yuan Shu, Hayato Nakanishi, Karen Smayra, Armin Farzad, Kaibo Hu, Qi Liu, Su Pan, Richard A. F. Dixon, Yue Wu, Peng Cai, Peng Yu, Pengyang Li

## Abstract

**Aims:** Cardiac arrhythmia is a rare complication after vaccination. Recently, reports of arrhythmia after COVID-19 vaccination have increased. Whether the risk for cardiac arrhythmia is higher with COVID-19 vaccines than with non–COVID-19 vaccines remains controversial. This meta-analysis explored the incidence of arrhythmia after COVID-19 vaccination and compared it with the incidence of arrhythmia after non–COVID-19 vaccination.

**Methods:** We searched the MEDLINE, Scopus, Cochrane Library, and Embase databases for English-language studies reporting the incidence of arrhythmia (the primary endpoint) after vaccination from January 1, 1947 to October 28, 2022. Secondary endpoints included incidence of tachyarrhythmia and all-cause mortality. Subgroup analyses were conducted to evaluate the incidence of arrhythmia by age (children [<18 years] versus adults [≥18 years]), vaccine type (mRNA COVID-19 vaccine versus non-mRNA COVID-19 vaccine; individual non–COVID-19 vaccines versus COVID-19 vaccine), and COVID-19 vaccine dose (first versus second versus third). Random-effects meta-analyses were performed, and the intrastudy risk for bias and the certainty of evidence were evaluated. This study was registered with PROSPERO (CRD42022365912).

**Results:** The overall incidence of arrhythmia from 36 studies (1,528,459,662 vaccine doses) was 291.8 (95% CI 111.6-762.7) cases per million doses. The incidence of arrhythmia was significantly higher after COVID-19 vaccination (2263.4 [875.4-5839.2] cases per million doses; 830,585,553 doses, 23 studies) than after non–COVID-19 vaccination (9.9 [1.3-75.5] cases per million doses; 697,874,109 doses, 14 studies; *P*<0.01). Compared with COVID-19 vaccines, the influenza, pertussis, human papillomavirus, and acellular pertussis vaccines were associated with a significantly lower incidence of arrhythmia.

The incidence of tachyarrhythmia was significantly higher after COVID-19 vaccination (4367.5 [1535.2-12,360.8] cases per million doses; 1,208,656 doses, 15 studies) than after non– COVID-19 vaccination (25.8 [4.5-149.4] cases per million doses; 179,822,553 doses, 11 studies; *P*<0.01). Arrhythmia was also more frequent after the third dose of COVID-19 vaccine (19,064.3 [5775.5-61,051.2] cases per million doses; 7968 doses, 3 studies) than after the first dose (3450.9 [988.2-11,977.6] cases per million doses; 41,714,762 doses, 12 studies; *P*=0.05) or second dose (2262.5 [2205.9-2320.7] cases per million doses; 34,540,749 doses, 10 studies; *P*<0.01). All-cause mortality was comparable between the COVID-19 and non–COVID-19 vaccination groups.

**Conclusions:** The overall risk for arrhythmia after COVID-19 vaccination was relatively low, although it was higher in COVID-19 vaccine recipients than in non–COVID-19 vaccine recipients. This increased risk should be evaluated along with other important factors, such as the incidence of local outbreaks and the risk for arrhythmia due to COVID infection itself, when weighing the safety and efficacy of COVID-19 vaccines.

## INTRODUCTION

As of October 2022, more than 12.79 billion COVID-19 vaccine doses have been administered globally.^1^ Because of the urgency of the pandemic, various COVID-19 vaccines were granted emergency approval before all three clinical trial phases could be completed.^2^ Although COVID-19 vaccines are generally considered safe, adverse events after vaccination, such as thromboembolisms, myocarditis, and arrhythmia, are increasingly being reported.^3-5^ Arrhythmias are a known but rare adverse reaction to viral vaccination and have also occurred after other vaccinations, including those for smallpox and human papillomavirus (HPV).^6-8^

The mRNA COVID-19 vaccines trigger innate immunity, cytotoxicity, and helper T cells, especially B cells, to promote the induction of neutralizing antibodies^9^; however, these reactions can also affect the cardiovascular system and cause serious adverse events, including arrhythmia.^10,11^ The Pfizer-BioNTech COVID-19 vaccine (BNT162B2) clinical trial reported a case of paroxysmal ventricular arrhythmia, but the exact mechanism was not found.^12^ Three cases of tachycardia after vaccination with the Pfizer BNT162B2 vaccine were reported in patients with a history of COVID-19 infection,^13^ and postural orthostatic tachycardia was observed in a healthy patient 6 days after the first dose of the Pfizer vaccine was administered.^14^ However, no consensus has been reached among clinicians as to whether these findings indicate true increased risk for arrhythmia or simply reflect recall bias.

To resolve this controversy, we performed a systematic review and meta-analysis of relevant observational studies to explore the risk for cardiac arrhythmia after COVID-19 and non–COVID-19 vaccination by characterizing the postvaccination incidence of cardiac arrhythmia. To our knowledge, this is the first meta-analysis to evaluate the safety and efficacy of COVID-19 versus non–COVID-19 vaccination within the context of cardiac arrhythmia.

## METHODS

### Search Strategy and Selection Criteria

This systematic review and meta-analysis was prospectively registered with PROSPERO (CRD42022365912) and conducted according to PRISMA (Preferred Reporting Items for Systematic Review and Meta-Analysis) guidelines^15^ (Supplemental Table S1).

The search strategy was designed and conducted by librarians with input from the study’s principal investigator. We comprehensively searched the MEDLINE, Scopus, Cochrane Library, and Embase databases for eligible observational literature from January 1, 1947 to October 28, 2022; search strategies are shown in Supplemental Table S2. Review articles and reference lists of the included studies also were retrieved as sources of grey literature. Observational studies of the general population who developed arrhythmia after vaccination were included. We excluded randomized controlled trials, case reports, case series, abstracts, review articles, nonhuman studies, non-article studies, non–English-language studies, and studies that did not report the number of doses.

### Data Extraction and Risk for Bias Assessment

Two researchers (XT, PX) independently retrieved relevant literature and extracted data in a predesigned extraction format (Supplemental Table S3). For repeated studies, we chose to include articles with the largest amount of information and the most complete data. The Joanna Briggs Institute (JBI) checklist was used to evaluate intrastudy risk for bias.^16^ To evaluate the certainty of evidence, we used the Grading of Recommendations Assessment, Development, and Evaluation (GRADE) system,^17^ which categorizes the quality of outcomes into four levels according to five metrics: risk for reporting bias, risk for publication bias, inconsistency, indirectness, and imprecision. Study assessment and selection were performed independently by two investigators (XT, PX). Any disputes were resolved by a third researcher (MH).

### Data Synthesis and Statistical Analyses

The primary outcome was the incidence of cardiac arrhythmia after any vaccination. Secondary outcomes were incidence of tachyarrhythmia and all-cause mortality after any vaccination. Because arrhythmia was defined differently across articles, we defined arrhythmia as including tachycardia, bradycardia, supraventricular arrhythmia, ventricular arrhythmia, ventricular tachycardia, ventricular fibrillation, atrial fibrillation, atrial flutter, atrial tachycardia, atrioventricular re-entrant tachycardia, atrioventricular nodal re-entrant tachycardia, supraventricular tachycardia, atrioventricular block, and heart block. We defined tachyarrhythmia as including all types of tachycardia, atrial fibrillation, and atrial flutter.

Statistical analyses were completed by using R version 4.1.3.^18^ We conducted random-effects meta-analyses and used pooled proportions (events per million vaccinations) and risk ratios (inverse variance method of DerSimonian and Laird)^19^ with 95% CIs to analyze the incidence of cardiac arrhythmia, tachyarrhythmia, and all-cause mortality after vaccination. We also used logit transformation to avoid any concerns raised by Freeman-Tukey double arcsine transformation in rare-events analysis.^20,21^ We then compared these incidences between COVID-19 vaccines and non–COVID-19 vaccines. Heterogeneity was evaluated by using the χ^2^ test and *I*^*2*^ statistic. An *I*^*2*^ value of 0%–25% indicates insignificant statistical heterogeneity, 26%–50% indicates low heterogeneity, and 51%–100% indicates high heterogeneity.^22^ A sensitivity analysis that excluded studies with high risk for bias (JBI score <7) assessed the impact of intrastudy risk for bias in reporting arrythmia. Publication bias was explored by using Egger’s linear regression tests and was visualized with funnel plots (Supplemental Figure S1); *P*<0.05 indicated strong publication bias.

We analyzed differences in the incidence of arrhythmia among prespecified subgroups, stratifying by: age (children [<18 years] versus adults [≥18 years]); vaccine type (mRNA COVID-19 vaccine versus non-mRNA COVID-19 vaccine, and COVID-19 vaccine versus individual non– COVID-19 vaccines, including HPV, acellular pertussis, influenza, smallpox, and mixed [epidemic cerebrospinal meningitis and pneumococcal conjugate vaccine]); and COVID-19 vaccine dose (first versus second versus third).

## RESULTS

### Study Selection and Study Characteristics

We identified 6,314 potentially eligible studies. After removing duplicates and filtering by title and abstract, 154 articles were left to evaluate by examining the full text. After applying our inclusion and exclusion criteria, 36 observational studies encompassing 1,528,459,662 vaccine doses were included in our meta-analysis (Figure 1).

**FIGURE 1.**
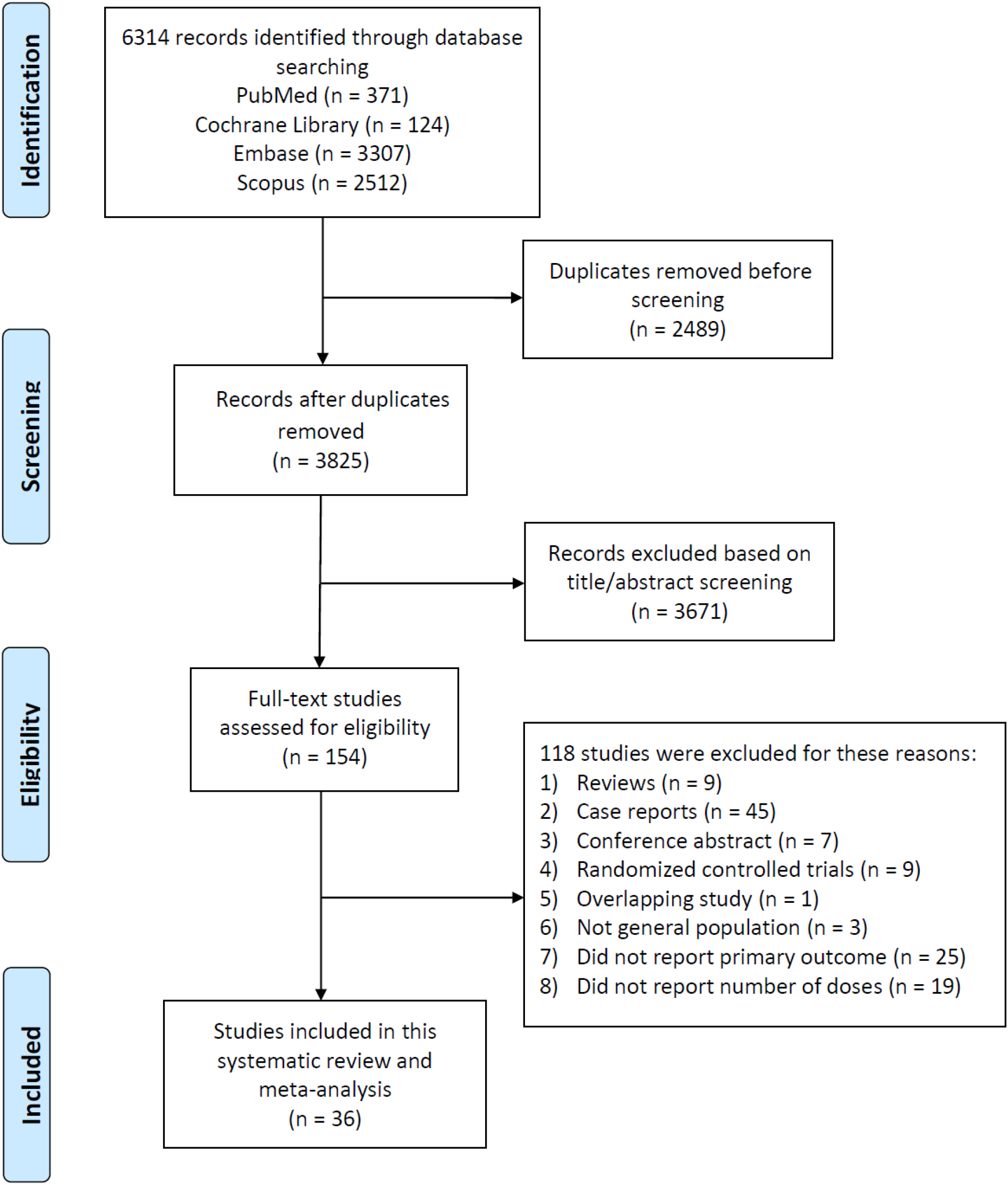
STUDY SELECTION.

Twenty-three studies reported on 830,585,553 doses of COVID-19 vaccines,^3,23-44^ 3 studies reported on 154,220,042 doses of HPV vaccines,^6,45,46^ 5 studies on 513,115,509 doses of influenza vaccines,^25,47-50^ 2 studies on 4,952,349 doses of acellular pertussis vaccines,^51,52^ 3 studies on 1,386,209 doses of smallpox vaccines,^50,53,54^ and 2 studies on 24,200,000 doses of mixed vaccines (epidemic cerebrospinal meningitis and pneumococcal conjugate vaccine).^55^ In the 23 studies involving COVID-19 vaccination, 647,308,174 doses of mRNA vaccines and 183,238,205 doses of non-mRNA vaccines were administered.

Basic characteristics of the articles included are summarized in Supplemental Table S4. The funnel plot and Egger’s test indicate no publication bias in this meta-analysis (Supplemental Figure S1). The JBI evaluation of intrastudy risk for bias and the GRADE assessment are provided in Supplemental Tables S5 and S6. Sensitivity analyses excluding studies with high risk for bias (JBI score >7) found no significant change in the pooled incidence of arrhythmia for COVID-19 and non–COVID-19 vaccines (Supplemental Table S7).

### Primary and Secondary Outcomes

The pooled results based on the random-effects model showed that the incidence of arrhythmia in the general population after vaccination was 291.8 (95% CI 111.6-762.7) cases per million vaccine doses; moderate certainty, Egger’s test *P*=0.92 (Figure 2; Supplemental Figure S1). The incidence of arrhythmia was significantly higher after COVID-19 vaccination (2263.4 [95% CI 875.4-5839.2] cases per million vaccine doses; low certainty) than after non– COVID-19 vaccination (9.9 [95% CI 1.3-75.5] cases per million vaccine doses; high certainty; *P*<0.01).

**FIGURE 2.**
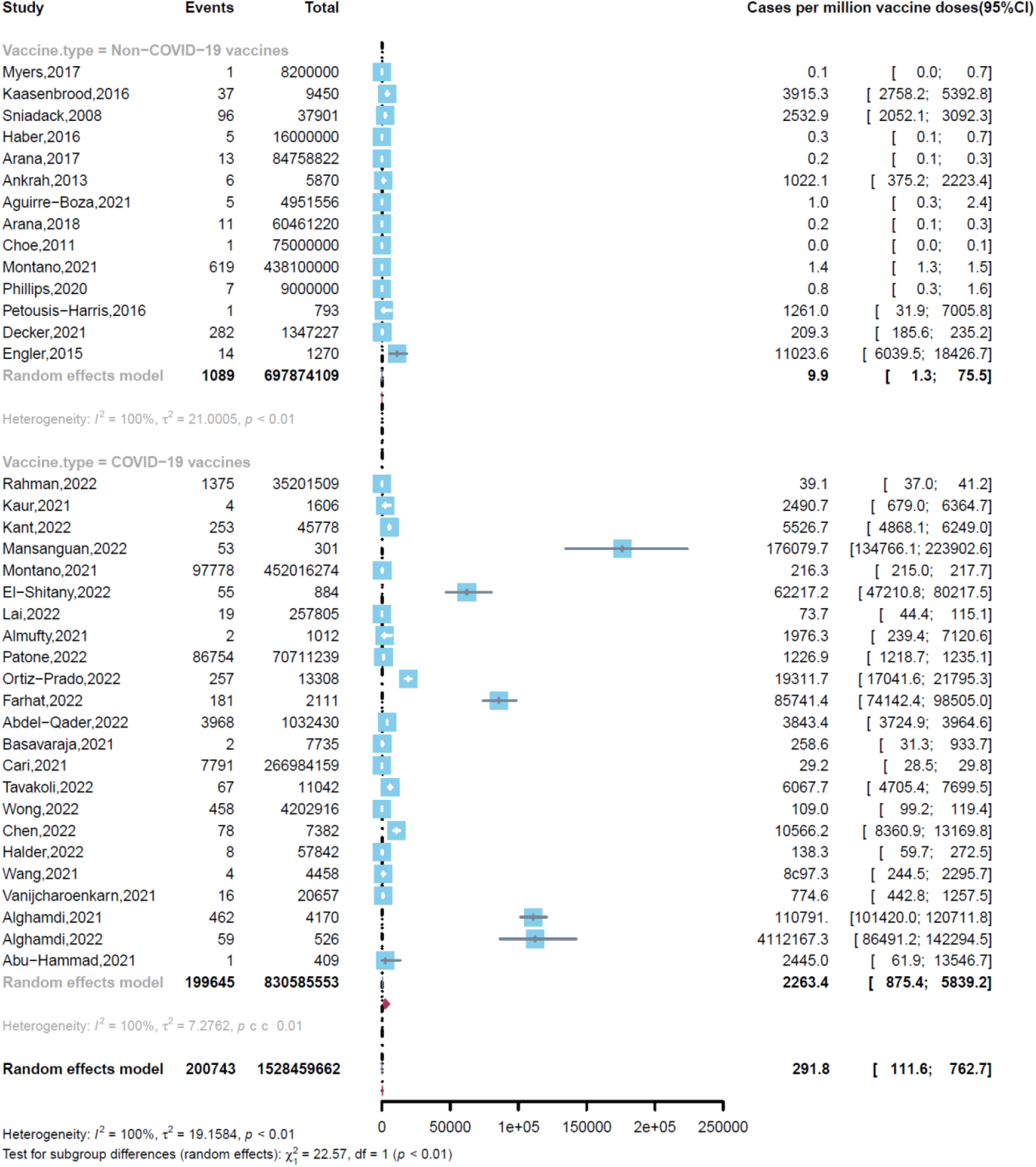
INCIDENCE OF ARRHYTHMIA IN THE GENERAL POPULATION AFTER VACCINATION.

The overall incidence of tachyarrhythmia was 487.1 (95% CI 127.3-1862.2) cases per million vaccine doses (Table 1, Supplemental Figure S2), of which 4367.5 (95% CI 1535.2-12,360.8) cases were COVID-19 vaccine recipients and 25.8 (95% CI 4.5-149.4) were cases were non–COVID-19 vaccine recipients; *P*<0.01.

**Table 1.** Subgroup analyses of the incidence of arrhythmia after COVID-19 and non-COVID-19 vaccination.

|  | **Study, n** | **Vaccine doses, n** | **Cardiac arrhythmia cases per million vaccine doses (95% CI)** | **P** |
| --- | --- | --- | --- | --- |
| Type of vaccine |  |  |  | <0.01 |
| Human papillomavirus | 3 | 154,220,042 | 0.3 (0.1-0.5) |  |
| COVID-19 | 23 | 830,585,553 | 2263.4 (875.4-5839.2) |  |
| Influenza | 5 | 513,115,509 | 53.2 (27.7-102.3) |  |
| Acellular pertussis | 2 | 4,952,349 | 21.1 (0.3-1739.9) |  |
| Smallpox | 3 | 1,386,209 | 1747.5 (275.9-10,982.9) |  |
| Mixed | 2 | 24,200,000 | 0.2 (0.1-0.6) |  |
| Tachyarrhythmia |  |  |  | <0.01 |
| Non-COVID-19 vaccines | 10 | 179,821,283 | 14.0 (3.4-58.2) |  |
| COVID-19 vaccines | 8 | 1,113,212 | 4518.0 (1549.4-13,099.3) |  |
| Dose |  |  |  | <0.01 |
| First | 12 | 41,714,762 | 3450.9 (988.2-11,977.6) |  |
| Second | 10 | 34,540,749 | 2262.5 (2205.9-2320.7) |  |
| Third | 3 | 7,968 | 19,064.3 (5775.5-61,051.2) |  |

Forest plots of the studies included in the subgroup analyses can be viewed in the
Supplemental Materials (Supplemental Figures S2, S4-S8, and S10).

The pooled all-cause mortality after vaccination was 2.1 (95% CI 0.5-9.0) events per million doses (126,033,203 doses, 8 studies; high certainty). No statistical difference was observed between COVID-19 vaccination (4.5 [95% CI 0.6-34.2] events per million doses; 40,436,855 doses, 3 studies) and non–COVID-19 vaccination (1.3 [95% CI 0.4-4.8] events per million doses; 85,596,348 doses, 5 studies; *P*=0.32) (Supplemental Figure S3).

### Subgroup Analyses

#### COVID-19 Vaccines versus Individual Non–COVID-19 Vaccines

We found significant differences in the incidence of arrhythmia between COVID-19 vaccines and individual non–COVID-19 vaccines; global *P*<0.01 (Table 1, Supplemental Figure S4). Compared with COVID-19 vaccination (2263.4 [95% CI 875.4-5839.2] cases per million doses), the incidence of arrhythmia was significantly lower after influenza vaccination (53.2 [95% CI 27.7-102.3] cases per million doses; *P*<0.01) (Supplemental Figure S5), HPV vaccination (0.3 [95% CI 0.1-0.5] cases per million doses; *P*<0.01) (Supplemental Figure S6), and acellular pertussis vaccination (21.1 [95% CI 0.3-1739.9] cases per million doses; *P*=0.04) (Supplemental Figure S7), whereas no difference was found after smallpox vaccination (1747.5 [95% CI 275.9-10,982.9] cases per million doses; *P*=0.81) (Supplemental Figure S8).

#### COVID-19 mRNA Vaccines versus COVID-19 Non-mRNA Vaccines

Among the 23 studies involving COVID-19 vaccination, no difference in the incidence of arrhythmia was found between the mRNA subgroup (1758.3 [95% CI 875.6-3527.7] cases per million doses; 647,308,174 doses, 18 studies) and non-mRNA subgroup (2528.1 [95% CI 960.9-6634.6] cases per million doses; 183,238,205 doses, 18 studies; *P*=0.55) (Supplemental Figure S9).

#### First versus Second versus Third Dose of COVID-19 Vaccines

The incidence of arrhythmia was significantly higher (global *P*<0.01) among those who received the third dose of COVID-19 vaccine (19,064.3 [95% CI 5775.5-61,051.2] cases per million doses; 7968 doses, 3 studies) than among those who received the first dose (3450.9 [988.2-11,977.6] cases per million doses; 41,714,762 doses, 12 studies; *P*=0.05) or the second dose (2262.5 [95% CI 2205.9-2320.7] cases per million doses; 34,540,749 doses, 10 studies; *P*<0.01) (Table 1, Supplemental Figure S10).

#### Adults versus Children

Subgroup analysis of recipient populations found no significant difference in the incidence of arrhythmia between adults (412.8 [95% CI 269.7-631.7] cases per million vaccine doses; 1,178,400,623 doses, 12 studies) and children (390.7 [95% CI 9.2–16,394.1] cases per million doses; 5,220,704 doses, 4 studies; *P*=0.98) (Supplemental Figure S11).

## DISCUSSION

The global rate of COVID-19 vaccination is increasing rapidly.^3-5^ Although vaccinations significantly protect us from viral infections, there is increasing evidence of various cardiovascular adverse events after vaccination for COVID-19, including cardiac arrythmias. Our systematic review and meta-analysis showed that the incidence of arrhythmia was significantly higher in people who received COVID-19 vaccines than in people who received non–COVID-19 vaccines (Figure 2), with the exception of smallpox vaccine. Additionally, the incidence of arrhythmia was comparable between COVID-19 mRNA and non-mRNA vaccines.

Before the COVID-19 pandemic, the global incidence of arrhythmia was largely attributed to age, sex, and socioeconomic status.^1,56^ Moreover, the incidence may be underestimated in select groups because of arrhythmia’s sometimes asymptomatic nature or a lack of routine screening.^57,58^ In the general population, the background incidence of arrhythmia is estimated to be 700-23,430 events per 1 million people.^1^ In our study, the incidence of arrhythmia after vaccination was 2263.4 (95% CI 875.4-5839.2) cases per million COVID-19 vaccine doses and 9.9 (95% CI 1.3-75.5) cases per million non–COVID-19 vaccine doses. These incidence levels fall within the background incidence range, suggesting a low risk for arrhythmia after taking any of the vaccines we pooled here.

Overall, the incidence of arrhythmia was significantly higher after COVID-19 vaccination than after non–COVID-19 vaccination. In the subgroup analyses, the incidence of arrhythmia was significantly lower after vaccination for influenza, pertussis, HPV, and acellular pertussis than after COVID-19 vaccination. The results of our study appear to be consistent with the literature: During the first 28 days after exposure to a COVID-19 vaccine, the risk for all cardiac arrhythmia subtypes increased.^3^ That said, our analysis differs from that in a recent case-control study of 884,828 individuals vaccinated with BNT162B2 in Israel,^59^ in which no increased risk for arrhythmia was found in the vaccination group compared with a control group. This contrast in findings might be explained by the different study populations and comparison groups, as we compared COVID-19 vaccines with non–COVID-19 vaccines. It is worth mentioning that COVID-19 vaccines are subject to more stringent monitoring than previous vaccines, given the rapid development of the current vaccine surveillance system. Therefore, arrhythmic events from traditional non-COVID-19 vaccines may have been relatively underreported.

Among people vaccinated for COVID-19, we found no significant difference in the incidence of arrhythmia between mRNA vaccine recipients and non-mRNA vaccine recipients. The incidence of arrhythmia did not differ between the first and second doses but was higher after the third dose compared with the previous two doses. A recent case series study, including 38,615,491 individuals vaccinated for COVID-19 and using a conditional Poisson regression model, found a lower risk for cardiac arrhythmia after the first and second doses of ChAdOx1 (adenovirus vector) and BNT162b2 (mRNA), along with a higher incidence of arrhythmia after a second dose of mRNA-1273.^3^ In our analysis, instead of considering individual vaccine types and doses as different variables, we clustered vaccines into two major categories, mRNA and non-mRNA, and we did not distinguish vaccine type when comparing the incidence of arrhythmia among different vaccine doses. The difference in analytical approach between these two studies may explain the discrepancy in their findings. Interestingly, the higher incidence of arrhythmia after a third dose of mRNA vaccination has not been reported in previous studies. The short-term local and systemic responses should be further explored.

The likelihood of new viral variants demands vaccination and boosters to defend ourselves during this ongoing pandemic.^60,61^ At the same time, the risks and benefits of vaccination should be assessed with caution when recommendations and policies are being debated, as weighted consideration must always be given to both the boosted immunity and potential adverse effects. Rare adverse events, including arrhythmia, may not become apparent in phase 3 clinical trials due to limited sample sizes, whereas postmarketing studies after the launch of a vaccine significantly augment the vaccination paradigm with more comprehensive understanding. Similar to the previous finding that younger males are more susceptible to myocarditis after COVID-19 vaccination,^62^ people with known risk factors for arrhythmia, including older age, male sex, white race, and cardiovascular comorbidities,^56^ should be appropriately addressed; these individuals may need to be monitored when receiving COVID-19 vaccinations. Nonetheless, the risk for such rare adverse events should be outweighed by the benefits of vaccination, including lower chance of infection, hospitalization, complications, and even death.^63,64^ These ideas would be crucial when drafting vaccination policies and should be dynamically updated as the pandemic evolves.

This study has several strengths. First, we aggregated reports of more than 1.5 billion vaccine doses—to our knowledge, the largest study to quantify the incidence of arrhythmia after vaccination. Second, we investigated whether COVID-19 vaccines were associated with a higher risk for arrhythmia compared with other traditional non–COVID-19 vaccines. Third, subgroup analysis of the vaccinated population by age, dose, and vaccine type could be used to identify potentially high-risk groups among vaccine recipients, help inform the public about the benefits and risks of receiving vaccines, and better balance the risk for adverse cardiovascular events against the risks of COVID-19 infection.

## Limitations

First, due to data limitations in the observational studies we included, we were unable to perform a subgroup analysis based on sex; in the subgroup analyses of age, we could only divide vaccine recipients into pediatric (<18 years) and adult (≥18 years) cohorts without further stratification. Second, there was heterogeneity in the definition of arrhythmia among the included studies. Self-reported arrhythmia was used in some studies, which could have been affected by subjective factors related to the patients themselves and may have led to biased data reporting of arrhythmic events. Third, the included studies were conducted for different time periods, which introduced heterogeneity and potential confounding factors into our analysis. Fourth, other types of non–COVID-19 vaccines were not included in this study, such as tetanus and hepatitis B vaccines, which may have resulted in underestimation of arrhythmia incidence in non–COVID-19 vaccine recipients. Fifth, all analyzed data were from retrospective studies, such that causal links in the results were weak, and there was a lack of long-term follow-up of patients. Finally, the burden and severity of the arrhythmias could not be analyzed.

## Conclusions

Our meta-analysis of more than 1.5 billion vaccine doses revealed that the incidence of arrhythmia was significantly higher after COVID-19 vaccination than after non–COVID-19 vaccination in general and after vaccination for influenza, pertussis, HPV, or acellular pertussis individually. Moreover, in the COVID-19 vaccination group, a higher incidence of arrhythmia was identified at the third dose compared with the previous two doses. The incidence of tachyarrhythmia and all-cause mortality after vaccination were similar for both COVID-19 and non–COVID-19 vaccine groups.

Our findings add new information about the safety profile of vaccines, especially COVID-19 vaccines. Although we found a greater risk for arrhythmia after COVID-19 vaccination than after non–COVID-19 vaccination, the incidence of local outbreaks and the risk for arrhythmia due to COVID-19 infection itself should be taken into consideration when evaluating the risks and benefits of COVID-19 vaccines.

## Data Availability

This paper uses publicly available data from published studies. No original data is available for sharing.

## ACKNOWLEDGEMENT

The authors wish to thank Jeanie F. Woodruff BS, ELS, of The Texas Heart Institute Department of Scientific Publications, for providing editorial support.

