## supplemental material for "Cardiac Arrhythmia after COVID-19 Vaccination versus Non–COVID-19 Vaccination: A Systematic Review and Meta-Analysis"

#### TABLE S1. PREFERRED REPORTING ITEMS FOR SYSTEMATIC REVIEWS AND META-ANALYSES (PRISMA) 2020 CHECKLIST

| Section and Topic | Item # | Checklist item | Location where item is reported |
|---|---|---|---|
| **TITLE** | | | |
| Title | 1 | Identify the report as a systematic review. | P1-2 |
| **ABSTRACT** | | | |
| Abstract | 2 | See the PRISMA 2020 for Abstracts checklist. | P3-4 |
| **INTRODUCTION** | | | |
| Rationale | 3 | Describe the rationale for the review in the context of existing knowledge. | P5 |
| Objectives | 4 | Provide an explicit statement of the objective(s) or question(s) the review addresses. | P5 |
| **METHODS** | | | |
| Eligibility criteria | 5 | Specify the inclusion and exclusion criteria for the review and how studies were grouped for the syntheses. | P6 |
| Information sources | 6 | Specify all databases, registers, websites, organisations, reference lists and other sources searched or consulted to identify studies. Specify the date when each source was last searched or consulted. | P6 |
| Search strategy | 7 | Present the full search strategies for all databases, registers and websites, including any filters and limits used. | P6, Table S2 |
| Selection process | 8 | Specify the methods used to decide whether a study met the inclusion criteria of the review, including how many reviewers screened each record and each report retrieved, whether they worked independently, and if applicable, details of automation tools used in the process. | P6 |
| Data collection process | 9 | Specify the methods used to collect data from reports, including how many reviewers collected data from each report, whether they worked independently, any processes for obtaining or confirming data from study investigators, and if applicable, details of automation tools used in the process. | P6 |
| Data items | 10a | List and define all outcomes for which data were sought. Specify whether all results that were compatible with each outcome domain in each study were sought (e.g. for all measures, time points, analyses), and if not, the methods used to decide which results to collect. | P7, Table S3 |

| Section and Topic | Item # | Checklist item | Location where item is reported |
|---|---|---|---|
| | 10b | List and define all other variables for which data were sought (e.g. participant and intervention characteristics, funding sources). Describe any assumptions made about any missing or unclear information. | P7, Table S3 |
| Study risk of bias assessment | 11 | Specify the methods used to assess risk of bias in the included studies, including details of the tool(s) used, how many reviewers assessed each study and whether they worked independently, and if applicable, details of automation tools used in the process. | P6-7 |
| Effect measures | 12 | Specify for each outcome the effect measure(s) (e.g. risk ratio, mean difference) used in the synthesis or presentation of results. | P7-8 |
| Synthesis methods | 13a | Describe the processes used to decide which studies were eligible for each synthesis (e.g. tabulating the study intervention characteristics and comparing against the planned groups for each synthesis (item #5)). | P7-8 |
| | 13b | Describe any methods required to prepare the data for presentation or synthesis, such as handling of missing summary statistics, or data conversions. | P7-8 |
| | 13c | Describe any methods used to tabulate or visually display results of individual studies and syntheses. | P7-8 |
| | 13d | Describe any methods used to synthesize results and provide a rationale for the choice(s). If meta-analysis was performed, describe the model(s), method(s) to identify the presence and extent of statistical heterogeneity, and software package(s) used. | P7-8 |
| | 13e | Describe any methods used to explore possible causes of heterogeneity among study results (e.g. subgroup analysis, meta-regression). | P7-8 |
| | 13f | Describe any sensitivity analyses conducted to assess robustness of the synthesized results. | P7-8 |
| Reporting bias assessment | 14 | Describe any methods used to assess risk of bias due to missing results in a synthesis (arising from reporting biases). | P7-8 |
| Certainty assessment | 15 | Describe any methods used to assess certainty (or confidence) in the body of evidence for an outcome. | P7-8 |

| Section and Topic | Item # | Checklist item | Location where item is reported |
|---|---|---|---|
| **RESULTS** | | | |
| Study selection | 16a | Describe the results of the search and selection process, from the number of records identified in the search to the number of studies included in the review, ideally using a flow diagram. | P8 |
| | 16b | Cite studies that might appear to meet the inclusion criteria, but which were excluded, and explain why they were excluded. | P8 |
| Study characteristics | 17 | Cite each included study and present its characteristics. | P8<br>Supplementary P11-19 |
| Risk of bias in studies | 18 | Present assessments of risk of bias for each included study. | Supplementary P20-22 |
| Results of individual studies | 19 | For all outcomes, present, for each study: (a) summary statistics for each group (where appropriate) and (b) an effect estimate and its precision (e.g. confidence/credible interval), ideally using structured tables or plots. | P8<br>Supplementary P11-19 |
| Results of syntheses | 20a | For each synthesis, briefly summarise the characteristics and risk of bias among contributing studies. | Table S6 |
| | 20b | Present results of all statistical syntheses conducted. If meta-analysis was done, present for each the summary estimate and its precision (e.g. confidence/credible interval) and measures of statistical heterogeneity. If comparing groups, describe the direction of the effect. | P8-10<br>Supplementary P27-36 |
| | 20c | Present results of all investigations of possible causes of heterogeneity among study results. | P8-10 |
| | 20d | Present results of all sensitivity analyses conducted to assess the robustness of the synthesized results. | P8-10 |
| Reporting biases | 21 | Present assessments of risk of bias due to missing results (arising from reporting biases) for each synthesis assessed. | Table S6 |
| Certainty of evidence | 22 | Present assessments of certainty (or confidence) in the body of evidence for each outcome assessed. | Table S6 |
| **DISCUSSION** | | | |
| Discussion | 23a | Provide a general interpretation of the results in the context of other evidence. | P10-13 |

| Section and Topic | Item # | Checklist item | Location where item is reported |
|---|---|---|---|
| | 23b | Discuss any limitations of the evidence included in the review. | P13 |
| | 23c | Discuss any limitations of the review processes used. | P13 |
| | 23d | Discuss implications of the results for practice, policy, and future research. | P13-14 |
| **OTHER INFORMATION** | | | |
| Registration and protocol | 24a | Provide registration information for the review, including register name and registration number, or state that the review was not registered. | P6 |
| | 24b | Indicate where the review protocol can be accessed, or state that a protocol was not prepared. | P6 |
| | 24c | Describe and explain any amendments to information provided at registration or in the protocol. | P6 |
| Support | 25 | Describe sources of financial or non-financial support for the review, and the role of the funders or sponsors in the review. | P14 |
| Competing interests | 26 | Declare any competing interests of review authors. | P14 |
| Availability of data, code and other materials | 27 | Report which of the following are publicly available and where they can be found: template data collection forms; data extracted from included studies; data used for all analyses; analytic code; any other materials used in the review. | P14 |

**TABLE S2. DETAILED DESCRIPTION OF THE SEARCH STRATEGY**

| | |
|---|---|
| **PubMed** | |
| #1 | Arrhythmias, Cardiac [MeSH Terms] |
| #2 | (dysrhythmia[Title/Abstract]) OR (arrhythmi*[Title/Abstract]) OR (dysrhythmi*[Title/Abstract]) OR (tachycardia[Title/Abstract]) OR bradycardia[Title/Abstract] OR (fibrillation[Title/Abstract]) OR (flutter[Title/Abstract]) OR (supraventricular arrhythmia[Title/Abstract]) OR (ventricular arrhythmia[Title/Abstract]) OR (ventricular tachycardia[Title/Abstract]) OR (ventricular fibrillation[Title/Abstract]) OR (atrial fibrillation[Title/Abstract]) OR (atrial flutter[Title/Abstract]) OR (atrial tachycardia[Title/Abstract]) OR (atrioventricular re-entrant tachycardia[Title/Abstract]) OR (atrioventricular nodal re-entrant tachycardia[Title/Abstract]) OR (supraventricular tachycardia[Title/Abstract]) OR (atrioventricular block[Title/Abstract]) OR (AV block[Title/Abstract]) OR (heart block[Title/Abstract]) |
| #3 | #1 OR #2 |
| #4 | vaccines [MeSH Terms] |
| #5 | vaccine[Title/Abstract]) |
| #6 | #4 OR #5 |
| #7 | #3 AND #6 |
| **Embase** | |
| #1 | Heart arrhythmia/exp |
| #2 | (dysrhythmia OR arrhythmi* OR dysrhythmi* OR tachycardia OR bradycardia OR fibrillation OR flutter OR supraventricular arrhythmia OR ventricular arrhythmia OR ventricular tachycardia OR ventricular fibrillation OR atrial fibrillation OR atrial flutter OR atrial tachycardia |

| | |
|---|---|
| | OR atrioventricular re-entrant tachycardia OR atrioventricular nodal re-entrant tachycardia OR supraventricular tachycardia OR atrioventricular block OR AV block OR heart block):ti,ab,kw |
| #3 | #1 OR #2 |
| #4 | vaccines/exp |
| #5 | vaccine:ti,ab,kw |
| #6 | #4 OR #5 |
| #7 | #3 AND #6 |
| **Cochrane Database of Systematic Reviews** | |
| #1 | MeSH descriptor:[Arrhythmias, Cardiac]explode all trees |
| #2 | (dysrhythmia OR arrhythmi* OR dysrhythmi* OR tachycardia OR bradycardia OR fibrillation OR flutter OR supraventricular arrhythmia OR ventricular arrhythmia OR ventricular tachycardia OR ventricular fibrillation OR atrial fibrillation OR atrial flutter OR atrial tachycardia OR atrioventricular re-entrant tachycardia OR atrioventricular nodal re-entrant tachycardia OR supraventricular tachycardia OR atrioventricular block OR AV block OR heart block):ti,ab,kw |
| #3 | #1 OR #2 |
| #4 | MeSH descriptor:[vaccines]explode all trees |
| #5 | vaccine:ti,ab,kw |
| #6 | #4 OR #5 |
| #7 | #3 AND #6 |

| **Scopus** | |
|---|---|
| #1 | TITLE-ABS-KEY(“arrhythmia” OR “dysrhythmia” OR “arrhythmi*” OR “dysrhythmi*” OR “tachycardia” OR “bradycardia” OR “fibrillation” OR “flutter” OR “supraventricular arrhythmia” OR “ventricular arrhythmia” OR “ventricular tachycardia” OR “ventricular fibrillation” OR “atrial fibrillation” OR “atrial flutter” OR “atrial tachycardia” OR “atrioventricular re-entrant tachycardia” OR “atrioventricular nodal re-entrant tachycardia” OR “supraventricular tachycardia” OR “atrioventricular block” OR “AV block” OR “heart block”) |
| #2 | TITLE-ABS-KEY(vaccines OR vaccine) |
| #3 | #1 AND #2 |

**TABLE S3. DATA COLLECTION TEMPLATE: CARDIAC ARRHYTHMIAS IN VACCINATION**

**Study characteristics:** Study authors, year published, vaccine characteristics, No. of doses, characteristics of those receiving vaccination, definitions of arrhythmia, outcomes reported, all mortality, follow-up duration.

**Baseline demographics:** Age, number or proportion of males, comorbidities

**Vaccine characteristics:** COVID-19 vaccine, HPV, acellular pertussis, influenza, smallpox, epidemic cerebrospinal meningitis and pneumococcal conjugate vaccine

**Outcomes:** arrhythmia, irregular heartbeat, atrial fibrillation, tachycardia, bradycardia

#### TABLE S4. DEMOGRAPHICS AND OUTCOMES OF INCLUDED STUDIES

| Study | Country | Vaccine characteristics | Number of doses | Characteristics of those receiving vaccination | Definitions of arrhythmia | Outcomes reported | All-cause mortality | Follow-up duration |
|---|---|---|---|---|---|---|---|---|
| Myers 2017 | USA | Menveo | 8200000 | Median: 12 y<br>Male 47% | Self-report (VAERS and MedDRA) | Postural orthostatic tachycardia syndrome: 1 | 1 | - |
| Kaasenbrood 2016 | Netherlands | Influenza | 9450 | 64.1±16.5 y<br>5075 males | ECG | Atrial fibrillation: 37 | - | - |
| Sniadack 2008 | USA | Smallpox | 37901 | Median: 48 y<br>Male 36% | Self-report (VAERS) | Arrhythmias: 7<br>Palpitations: 89 | 2 | 5–12 mo |
| Haber 2016 | USA | Pneumococcal conjugate vaccine (PCV13) | 16000000 | ≥19 y | Self-report (VAERS) | Atrial fibrillation: 4 | ≥65 y: 14 | - |
| Arana 2017 | USA | HPV | 80138237 4vHP<br>715000 2vHPV<br>3905585 9vHPV | Children and adults | Clinically confirmed (diagnostic criteria for POTS) | Postural orthostatic tachycardia syndrome:<br>4vHP: 13<br>2vHPV: 0<br>9vHPV: 0 | - | - |
| Ankrah 2013 | Netherlands | A(H1N1) 2009 | 5870 | Mean: 34 y<br>2111 males | Self-report (MedDRA) | Tachycardia: 6 | - | 1 wk |
| Aguirre-Boza 2021 | Chile | Acellular pertussis | 4951556 | Infants <2 y | Self-report (MedDRA) | Bradycardia: 5 | - | - |
| Arana 2018 | USA | 4vHPV | 60461220 | Children and adults | Clinically confirmed (confirmed cases following clinical review of reports) | Postural orthostatic tachycardia syndrome: 11 | 92 | - |
| Choe 2011 | Korea | Trivalent inactivated influenza | 75000000 | Children and adults | Self-report (KVICP) | Bradycardia: 1 | - | - |

| Study | Country | Vaccine characteristics | Number of doses | Characteristics of those receiving vaccination | Definitions of arrhythmia | Outcomes reported | All-cause mortality | Follow-up duration |
|---|---|---|---|---|---|---|---|---|
| Montano 2021 | Germany | Influenza | 438100000 | - | Self-report (VAERS and EudraVigilance) | Arrhythmia: 619 | - | - |
| | | COVID-19: AstraZeneca or Pfizer or Moderna or Janssen | 34643783 AstraZeneca 32233117 Janssen 105518547 Moderna 279620827 Pfizer | Age 18-64 y: 335766004 ≥65 y: 116250604<br><br>226008303.5 males | | Arrhythmia: 97778 AstraZeneca: 20614 Janssen: 5574 Moderna: 23869 Pfizer: 47721 | - | |
| Phillips 2020 | Australia | 4vHPV | 9000000 | Children and adult | Self-report (VAERS and MedDRA) | Postural orthostatic tachycardia syndrome: 7 | - | - |
| Petousis-Harris 2016 | New Zealand | Tdap | 793 | Mean: 32 y <20 y: 12 20-24 y: 65 25-29 y: 180 30-34 y: 307 35-39 y: 180 ≥40 y: 49<br><br>Women in wk 28–38 of pregnancy | - | Tachycardia: 1 | | 4 wk |
| Ab Rahman 2022 | Malaysia | COVID-19: BNT162b2 | 15387585 | | ICD-10 | | 8 | - |
| | | COVID-19: CoronaVac | 17030243 | | | | | |

| Study | Country | Vaccine characteristics | Number of doses | Characteristics of those receiving vaccination | Definitions of arrhythmia | Outcomes reported | All-cause mortality | Follow-up duration |
|---|---|---|---|---|---|---|---|---|
| | | COVID-19: ChAdOx1 | 2744507 | Age<br>12-17 y: 17356<br>18-39 y: 10247762<br>40-59 y: 6263483<br>60+ y: 3278612<br><br>9919733 males | | Arrhythmias: 1375<br>(834 BNT162b2; 485 CoronaVac; 53 ChAdOx1; 3 Others) | | |
| | | COVID-19: Others | 39174 | | | | | |
| Kaur 2021 | India | COVID-19: ChAdOx1 | 1st dose: 804<br>2nd dose: 802 | Mean (±SD): 38.44 (±11.47)<br>573 males | Self-report (MedDRA) | Tachycardia:1st dose: 4<br>2nd dose: 0 | - | 15 mo |
| Kant 2022 | Netherlands | COVID-19: AstraZeneca | 14338 | Age<br>12-20 y: 369<br>21-65 y: 17527<br>66-80 y: 6757<br>>80 y: 2901<br><br>10622 males | Self-report (MedDRA) | Arrhythmia: 14<br>(6 AstraZeneca; 7 Pfizer; 1 Moderna; 0 Janssen)<br>Tachycardia: 14<br>(8 AstraZeneca; 1 Pfizer; 4 Moderna; 1 Janssen)<br>Atrial fibrillation: 11<br>(4 AstraZeneca; 6 Pfizer; 1 Moderna; 0 Janssen)<br>Heart rate irregular: 10<br>(2 AstraZeneca; 6 Pfizer; 1 Moderna; 1 Janssen)<br>Atrioventricular block complete: 1<br>(0 AstraZeneca; 1 Pfizer; 0 Moderna; 0 Janssen) | - | 6 mo |
| | | COVID-19: Pfizer | 23115 | | | | | |
| | | COVID-19: Moderna | 5867 | | | | | |
| | | COVID-19: Janssen | 2458 | | | | | |

| Study | Country | Vaccine characteristics | Number of doses | Characteristics of those receiving vaccination | Definitions of arrhythmia | Outcomes reported | All-cause mortality | Follow-up duration |
|---|---|---|---|---|---|---|---|---|
| | | | | | | Palpitation: 203 (106 AstraZeneca; 48 Pfizer; 32 Moderna; 17 Janssen) | | |
| Mansanguan 2022 | Thailand | COVID-19: BNT162b2 | 301 | Age<br>Mean (±SD): 15 (±1.6)<br>13-15 y: 207<br>16-18 y: 94<br><br>202 males (adolescents) | ECG | Sinus rhythm with sinus arrhythmia: 22<br>Sinus tachycardia: 20<br>Sinus bradycardia: 4<br>Premature atrial contraction: 3<br>Premature ventricular contraction: 2<br>Junctional escape rhythm: 1<br>Ectopic atrial rhythm: 1 | - | 14 d |
| El-Shitany 2022 | Egypt | COVID-19: Pfizer | 2nd dose: 442<br>3rd dose: 442 | Age<br>< 60 y: 367<br>≥ 60 y: 75<br><br>123 males | Self-report (a Google form-based questionnaire, the scientific terms for the symptoms were written and explained in the public language) | Arrhythmia: 2nd dose: 25<br>3rd dose: 30 | - | - |
| Farhat 2022 | Saudi Arabia | COVID-19: Pfizer | 1st dose: 772<br>2nd dose: 816<br>3rd dose: 142 | Age<br>12-17 y: 33<br>18-29 y: 239 | Self-report (a Google form-based questionnaire, the | Fast or irregular heartbeat:<br>1st dose: 95<br>2nd dose: 85 | - | - |

| Study | Country | Vaccine characteristics | Number of doses | Characteristics of those receiving vaccination | Definitions of arrhythmia | Outcomes reported | All-cause mortality | Follow-up duration |
|---|---|---|---|---|---|---|---|---|
| | | COVID-19: Oxford | 1st dose: 217<br>2nd dose: 162<br>3rd dose: 2 | 30-49 y: 424<br>50-65 y: 301<br>>65 y: 21<br><br>390 males | scientific terms for the symptoms were written and explained in the public language) | 3rd dose: 1 | | |
| Lai 2022 | Hong Kong, China | COVID-19: BNT162b2 | 1st dose: 138141<br>2nd dose: 119664 | 12-18 y<br><br>1st dose: 69572 males<br>2nd dose: 61168 males | ICD-9 | Arrhythmia:<br>1st dose: 7<br>2nd dose: 12 | - | - |
| Almufty 2021 | Iraq | COVID-19: AstraZeneca | 608 | Age<br>18-30 y: 420<br>31-49 y: 424<br>50-69 y: 150<br>≥70 y: 18<br><br>609 males | Self-report (a standardized questionnaire platform was used to collect information) | Tachycardia:2<br>(1 AstraZeneca; 1 Pfizer) | - | - |
| | | COVID-19: Pfizer | 296 | | | | | |
| | | COVID-19: Sinopharm | 108 | | | | | |
| Patone 2022 | UK | COVID-19: ChAdOx1 | 1st dose: 20615911<br>2nd dose: 19754224 | 1st dose: Age<br>16-29 y: 5767025<br>30-39 y: 5910138<br>40+ y: 26938328<br>2nd dose: Age<br>16-29 y: 2258383<br>30-39 y: 4130788<br>40+ y: 25706577 | Clinically confirmed | Arrhythmia:<br>1st dose: 42740<br>2nd dose: 44014 | - | - |
| | | COVID-19: BNt162b2 | 1st dose: 16993389<br>2nd dose: 11972733 | | | | | |

| Study | Country | Vaccine characteristics | Number of doses | Characteristics of those receiving vaccination | Definitions of arrhythmia | Outcomes reported | All-cause mortality | Follow-up duration |
|---|---|---|---|---|---|---|---|---|
| | | COVID-19: mRNA-1273 | 1st dose: 1006191<br>2nd dose: 368791 | 1st dose: 12883163 males<br>2nd dose: 10905154 males | | | | |
| Ortiz-Prado 2022 | Ecuador | COVID-19: Pfizer | 1st dose: 2069<br>2nd dose:2069 | Age<br>10-20 y: 204<br>21-30 y: 1553<br>31-40 y: 1895<br>41-50 y: 1471<br>51-60 y: 997<br>61-70 y: 428<br>71-80 y: 82<br>81-90 y: 19<br>91-100 y: 5<br><br>2631 males | Self-report (online questionnaire) | Tachycardia: 257 (106 Pfizer; 101 AstraZeneca; 50 Sinovac) | - | - |
| | | COVID-19: AstraZeneca | 1st dose: 2553<br>2nd dose:2553 | | | | | |
| | | COVID-19: Sinovac | 1st dose: 2032<br>2nd dose:2032 | | | | | |
| Abdel-Qader 2022 | Jordan | COVID-19: BNT162b2 | 610591 | Mean: 36.5 y<br><br>622844 males | Self-report (telephone survey) | Tachycardia: 1st dose: 2430<br>2nd dose: 1538 | 11 | 8 mo |
| | | COVID-19: BBIBP-CorV | 279606 | | | | | |
| | | COVID-19: ChAdOx1 | 140843 | | | | | |
| | | COVID-19: Sputnik V | 1390 | | | | | |
| Basavaraja 2021 | India | COVID-19: COVISHIELD | 9292 | | Self-report (MedDRA) | Tachycardia: 1st dose: 2<br>2nd dose: 0 | - | 90 d |

| Study | Country | Vaccine characteristics | Number of doses | Characteristics of those receiving vaccination | Definitions of arrhythmia | Outcomes reported | All-cause mortality | Follow-up duration |
|---|---|---|---|---|---|---|---|---|
| | | COVID-19: COVAXIN | 2364 | ≥18 y | | | | |
| Cari 2021 | Italy | COVID-19: ChAdOx1 | 48071531 | ≥18 y | EudraVigilance European database | Tachycardia:2 (2918 ChAdOx1; 144 Ad26.COV2.S; 4729 BNT162b2) | - | - |
| | | COVID-19: Ad26.COV2.S | 5891506 | | | | | |
| | | COVID-19: BNT162b2 | 213021122 | | | | | |
| Decker 2021 | USA | Smallpox: ACAM2000 | 897227 | ≥17 y | ICD-9 and ICD-10 | Arrhythmia: 131 (10 ACAM2000; 121 Dryvax) | 1 | - |
| | | Smallpox: Dryvax | 450000 | | | Atrial Fibrillation: 1 (1 ACAM2000; 0 Dryvax) Supraventricular Tachycardia: 3 (3 ACAM2000; 0 Dryvax) Tachycardia Paroxysmal: 1 (1 ACAM2000; 0 Dryvax) Palpitation: 146 (28 ACAM2000; 118 Dryvax) | - | |
| Tavakoli 2022 | Iran | COVID-19: Soberana | 1315 | Mean: 14.5 y | Self-report (telephone survey) | Arrhythmia: 18 (4 Soberana; 14 Sinopharm) | - | - |
| | | COVID-19: Sinopharm | 9727 | 5374 males | | Palpitation: 49 | | |

| Study | Country | Vaccine characteristics | Number of doses | Characteristics of those receiving vaccination | Definitions of arrhythmia | Outcomes reported | All-cause mortality | Follow-up duration |
|---|---|---|---|---|---|---|---|---|
| | | | | | | (7 Soberana; 42 Sinopharm) | | |
| Wong 2022 | Hong Kong, China | COVID-19: BNT162b2 | 1st dose: 1308820<br>2nd dose: 1116677 | Mean: 49.8 y<br><br>1024086 males | ICD-9-CM | Arrhythmia: 458 (224 BNT162b2; 14 CoronaVac) | 1st dose: 42<br>2nd dose: 34 | - |
| | | COVID-19: CoronaVac | 1st dose: 955859<br>2nd dose: 821560 | | | | 1st dose: 53<br>2nd dose: 33 | |
| Abu-Hammad 2021 | Jordan | COVID-19: SinoPharm | 89 | Mean: 35.0 y<br><br>120 males | Self-report (online questionnaire) | Palpitations: 1 (0 SinoPharm; 0 Pfizer BioNTech; 1 AstraZeneca Vaxezevira) | - | - |
| | | COVID-19: Pfizer BioNTech | 141 | | | | | |
| | | COVID-19: AstraZeneca Vaxezevira | 179 | | | | | |
| Alghamdi 2021 | Saudi Arabia | COVID-19: BNT162b2 | 2601 | Age<br><20 y: 671<br>20-30 y: 1965<br>31-40 y: 671<br>41-50 y: 582<br>51-60 y: 227<br>>60 y: 54<br><br>1296 males | Self-report (online questionnaire) | Palpitations: 462 (193 BNT162b2; 269 ChAdOx1) | - | - |
| | | COVID-19: ChAdOx1 | 1569 | | | | | |

| Study | Country | Vaccine characteristics | Number of doses | Characteristics of those receiving vaccination | Definitions of arrhythmia | Outcomes reported | All-cause mortality | Follow-up duration |
|---|---|---|---|---|---|---|---|---|
| Alghamdi 2022 | Saudi Arabia | COVID-19: AstraZeneca | 526 | Mean: 39.7 y<br><br>244 males | Self-report (online questionnaire) | Palpitations: 59 | - | - |
| Chen 2022 | Taiwan, China | COVID-19: mRNA1273 | 5374 | Age<br><40 y: 3011<br><65 y: 3751<br>≥65 y: 620<br><br>2461 males | Self-report (VAERS) | Palpitations: 78 (46 mRNA1273; 24 BNT162b2; 8 MVC-COV1901 ) | - | 3 mo |
| | | COVID-19: BNT162b2 | 1182 | | | | | |
| | | COVID-19: MVC-COV1901 | 826 | | | | | |
| Engler 2015 | USA | Smallpox | 1st dose: 1081 | Mean: 25.3 y<br><br>1058 males | Self-report (telephone and/or electronic mail) | Palpitations: 12 | - | 58 mo |
| | | Influenza | 1st dose: 189 | | | Palpitations: 2 | | |
| Halder 2022 | Australia | COVID-19: BNT162b2 | 57,842 | Mean: 40.6 y<br><br>- | Self-report (electronic medical records) | Palpitations: 8 | - | - |
| Vanijcharoenkarn 2021 | USA | SARSCoV-2 mRNA Vaccines | 1st dose: 20,657 | - | Self-report (occupational health records) | Palpitations: 16 | - | - |
| Wang 2021 | China | COVID-19 vaccine：Aikewei | 4458 | - | Self-report (recorded in Children’s Hospital of Fudan University) | Palpitations: 4 | - | - |

**TABLE S5. JOANNA BRIGGS INSTITUTE CHECKLIST FOR PREVALENCE STUDIES**

| Study | 1 | 2 | 3 | 4 | 5 | 6 | 7 | 8 | 9 | Overall |
|---|---|---|---|---|---|---|---|---|---|---|
| Myers 2017 | √ | √ | √ | √ | √ | ? | √ | √ | ? | 7 |
| Kaasenbrood 2016 | √ | √ | √ | √ | √ | √ | √ | √ | √ | 9 |
| Sniadack 2008 | √ | √ | √ | ? | √ | × | √ | √ | √ | 7 |
| Haber 2016 | √ | √ | √ | √ | √ | × | × | √ | √ | 7 |
| Arana 2017 | √ | √ | √ | √ | √ | √ | √ | √ | √ | 9 |
| Ankrah 2013 | √ | √ | √ | √ | √ | × | × | √ | √ | 7 |
| Aguirre-Boza 2021 | √ | √ | √ | √ | √ | × | × | √ | √ | 7 |
| Arana 2018 | √ | √ | √ | √ | √ | √ | √ | √ | √ | 9 |
| Choe 2011 | √ | ? | √ | ? | √ | × | ? | √ | √ | 5 |
| Montano 2021 | √ | √ | √ | √ | √ | × | × | √ | √ | 7 |
| Phillips 2020 | √ | √ | √ | √ | √ | × | × | √ | √ | 7 |
| Petousis-Harris 2016 | √ | √ | × | √ | √ | ? | ? | √ | √ | 6 |
| Ab Rahman 2022 | √ | ? | √ | √ | √ | √ | √ | √ | √ | 8 |
| Kaur 2021 | √ | √ | √ | √ | √ | ? | ? | √ | √ | 7 |
| Kant 2022 | √ | √ | √ | √ | √ | × | √ | √ | √ | 8 |
| Mansanguan 2022 | √ | √ | × | √ | √ | √ | √ | √ | √ | 8 |
| El-Shitany 2022 | × | √ | × | √ | √ | × | √ | √ | √ | 6 |
| Farhat 2022 | √ | √ | √ | √ | √ | × | ? | √ | √ | 7 |
| Lai 2022 | √ | √ | √ | √ | √ | √ | √ | √ | √ | 9 |
| Almufty 2021 | √ | ? | √ | √ | √ | × | √ | √ | √ | 7 |
| Patone 2022 | √ | √ | √ | √ | √ | √ | √ | √ | √ | 9 |
| Ortiz-Prado 2022 | √ | √ | √ | √ | √ | × | √ | √ | √ | 8 |
| Abdel-Qader 2022 | √ | √ | √ | √ | √ | × | √ | √ | √ | 8 |

| | | | | | | | | | | |
|---|---|---|---|---|---|---|---|---|---|---|
| Basavaraja 2021 | √ | √ | √ | ? | √ | × | √ | √ | √ | 7 |
| Cari 2021 | √ | √ | √ | √ | √ | ? | ? | √ | √ | 7 |
| Decker 2021 | √ | √ | √ | ? | √ | √ | √ | ? | √ | 7 |
| Tavakoli 2022 | √ | √ | √ | √ | √ | × | √ | √ | √ | 8 |
| Wong 2022 | √ | √ | √ | √ | √ | √ | √ | √ | √ | 9 |
| Abu-Hammad 2021 | √ | √ | × | √ | √ | × | √ | √ | √ | 7 |
| Alghamdi 2021 | √ | ? | √ | √ | √ | × | √ | √ | √ | 7 |
| Alghamdi 2022 | × | √ | × | √ | √ | × | ? | √ | √ | 5 |
| Chen 2022 | × | √ | √ | √ | √ | × | √ | √ | √ | 7 |
| Engler 2015 | √ | √ | √ | √ | √ | × | √ | √ | √ | 8 |
| Halder 2022 | √ | √ | √ | √ | √ | × | × | ? | √ | 6 |
| Vanijcharoenkarn 2021 | √ | √ | √ | √ | √ | × | √ | ? | √ | 7 |
| Wang 2021 | √ | √ | √ | √ | √ | × | √ | ? | √ | 7 |

√ : Yes, ×: no, ?: unclear.

**Domains:**

1: Was the sample frame appropriate to address the target population?
2: Were study participants sampled in an appropriate way?
3: Was the sample size adequate?
4: Were the study subjects and the setting described in detail
5: Was the data analysis conducted with sufficient coverage of the identified sample?
6: Were valid methods used for the identification of the condition?
7: Was the condition measured in a standard, reliable way for all participants?
8: Was there appropriate statistical analysis?
9: Was the response rate adequate, and if not, was the low response rate managed appropriately?

#### TABLE S6. GRADING OF RECOMMENDATIONS, ASSESSMENT, DEVELOPMENT, AND EVALUATIONS

| Number of studies | Certainty assessment | | | | | | Effect | | Certainty | Importance |
|---|---|---|---|---|---|---|---|---|---|---|
| | **Study design** | **Risk for bias** | **Inconsistency** | **Indirectness** | **Imprecision** | **Other considerations** | **Number of individuals** | **Rate (95% CI)** | | |
| Incidence of cardiac arrhythmias (assessed with: per million vaccines) | | | | | | | | | | |
| 36 | Observational studies | Not serious | Serious[a] | Not serious | Not serious | None | 1528459662 | 15014.69 per million (3958.84-26,070.55) | ⨁⨁⨁◯ Moderate | CRITICAL |
| Incidence of cardiac arrhythmias in COVID-19 vaccines (assessed with: per million vaccines) | | | | | | | | | | |
| 23 | Observational studies | Not serious | Serious[a] | Not serious | Serious[b] | None | 830585553 | 24420.82 per million (6284.27-42,557.37) | ⨁⨁◯◯ Low | CRITICAL |
| Incidence of cardiac arrhythmias in non-COVID-19 vaccines (assessed with: per million vaccines) | | | | | | | | | | |
| 14 | Observational studies | Not serious | Not serious[c] | Not serious | Not serious | None | 697874109 | 867.56 per million (0.00-1867.84) | ⨁⨁⨁⨁ High | CRITICAL |
| Mortality (assessed with: per million vaccines) | | | | | | | | | | |
| 8 | Observational studies | Not serious | Not serious | Not serious | Not serious[d] | None | 126033203 | 8.15 per million (0.00-18.40) | ⨁⨁⨁⨁ High | CRITICAL |

**Explanations**

[a] Point estimates were sparsely distributed, and confidence intervals overlapped only occasionally.

[b] Compared with the combined estimation, the difference in confidence intervals is very large.

[c] Apart from one study, most of the point estimates were closely clustered, and the confidence intervals overlapped most of the time. Thus, we did not rate down for inconsistency.

[d] Compared with the combined estimation, the difference in confidence intervals is not very large. Thus, we did not rate down for imprecision.

**TABLE S7: RESULTS OF THE SENSITIVITY ANALYSES FOR THE PRIMARY META-ANALYSIS.**

| Sensitivity analysis | Pooled estimate | 95% CI |
|---|---|---|
| Excluding studies with high risk for bias | 277.8 arrhythmia cases per million vaccine doses | 150.2–513.7 |

**FIGURE S1. FUNNEL PLOT AND EGGER TEST FOR PRIMARY META-ANALYSIS OF CARDIAC ARRHYTHMIAS AFTER VACCINATION**

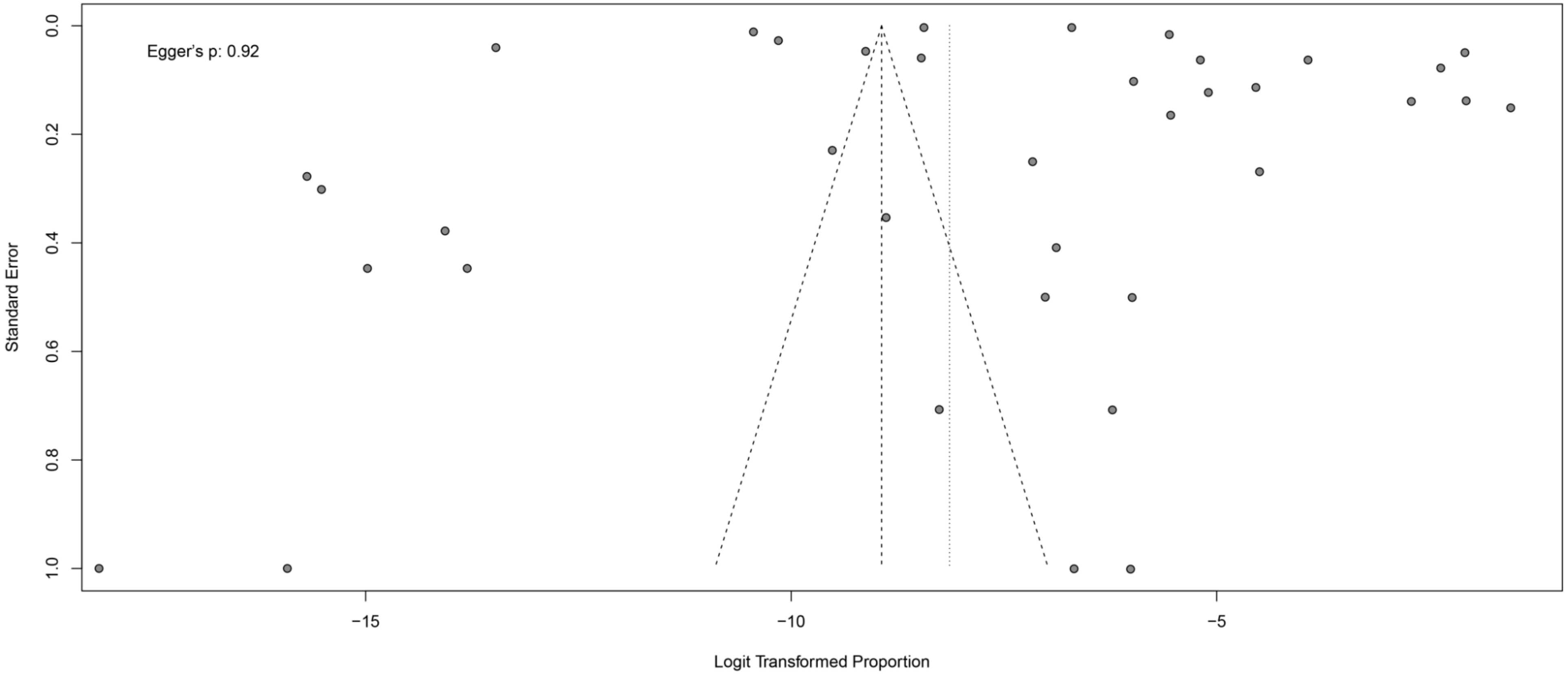

**FIGURE S2. INCIDENCE OF TACHYARRHYTHMIA AMONG THOSE RECEIVING COVID-19 AND NON–COVID-19 VACCINES**

| Study | Events | Total | Cases per million vaccine doses(95%CI) | |
|---|---|---|---|---|
| **Vaccine.type = Non−COVID−19 vaccines** | | | | |
| Myers,2017 | 1 | 8200000 | 0.1 | [ 0.0; 0.7] |
| Arana,2017 | 13 | 84758822 | 0.2 | [ 0.1; 0.3] |
| Ankrah,2013 | 6 | 5870 | 1022.1 | [ 375.2; 2223.4] |
| Arana,2018 | 11 | 60461220 | 0.2 | [ 0.1; 0.3] |
| Phillips,2020 | 7 | 9000000 | 0.8 | [ 0.3; 1.6] |
| Petousis−Harris,2016 | 1 | 793 | 1261.0 | [ 31.9; 7005.8] |
| Kaasenbrood,2016 | 37 | 9450 | 3915.3 | [ 2758.2; 5392.8] |
| Sniadack,2008 | 89 | 37901 | 2348.2 | [ 1886.2; 2888.9] |
| Haber,2016 | 5 | 16000000 | 0.3 | [ 0.1; 0.7] |
| Decker,2021 | 151 | 1347227 | 112.1 | [ 94.9; 131.5] |
| Engler,2015 | 14 | 1270 | 11023.6 | [ 6039.5; 18426.7] |
| Random effects model | **335** | **179822553** | **25.8** | **[ 4.5; 149.4]** |
| Heterogeneity: $I^2$ = 100%, $\tau^2$ = 20.3363, $p$ < 0.01 | | | | |
| **Vaccine.type = COVID−19 vaccines** | | | | |
| Kaur,2021 | 4 | 1606 | 2490.7 | [ 679.0; 6364.7] |
| Kant,2022 | 228 | 45778 | 4980.6 | [ 4356.3; 5668.8] |
| Mansanguan,2022 | 20 | 301 | 66445.2 | [ 41055.6; 100756.0] |
| Almufty,2021 | 2 | 1012 | 1976.3 | [ 239.4; 7120.6] |
| Ortiz−Prado,2022 | 257 | 13308 | 19311.7 | [ 17041.6; 21795.3] |
| Abdel−Qader,2022 | 3968 | 1032430 | 3843.4 | [ 3724.9; 3964.6] |
| Basavaraja,2021 | 2 | 7735 | 258.6 | [ 31.3; 933.7] |
| Tavakoli,2022 | 49 | 11042 | 4437.6 | [ 3284.7; 5862.6] |
| Chen,2022 | 78 | 7382 | 10566.2 | [ 8360.9; 13169.8] |
| Halder,2022 | 8 | 57842 | 138.3 | [ 59.7; 272.5] |
| Wang,2021 | 4 | 4458 | 897.3 | [ 244.5; 2295.7] |
| Vanijcharoenkarn,2021 | 16 | 20657 | 774.6 | [ 442.8; 1257.5] |
| Alghamdi,2021 | 462 | 4170 | 110791.4 | [101420.0; 120711.8] |
| Alghamdi,2022 | 59 | 526 | 112167.3 | [ 86491.2; 142294.5] |
| Abu−Hammad,2021 | 1 | 409 | 2445.0 | [ 61.9; 13546.7] |
| Random effects model | **5158** | **1208656** | **4367.5** | **[ 1535.2; 12360.8]** |
| Heterogeneity: $I^2$ = 100%, $\tau^2$ = 4.1194, $p$ < 0.01 | | | | |
| **Random effects model** | **5493** | **181031209** | **487.1** | **[ 127.3; 1862.2]** |

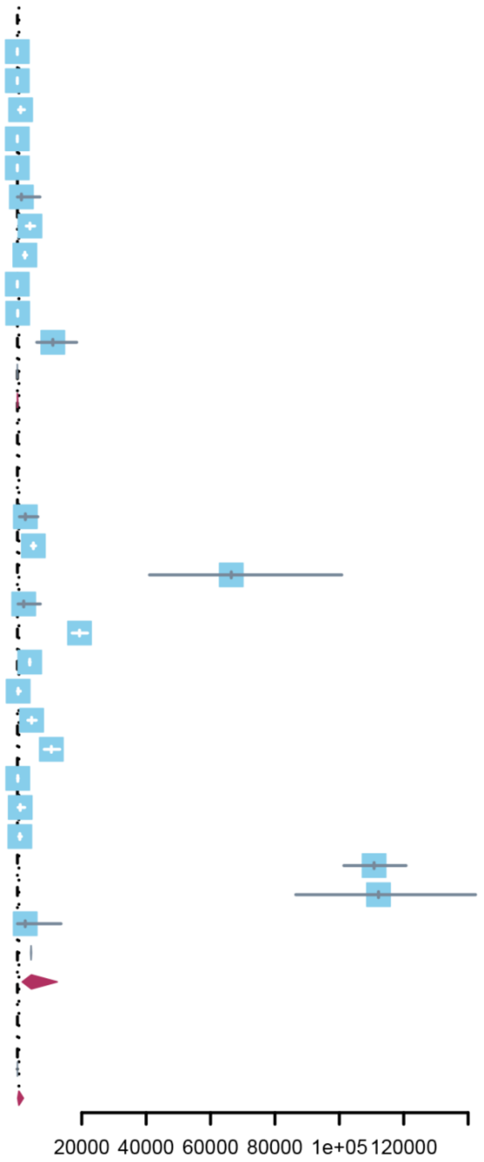

Heterogeneity: $I^2$ = 100%, $\tau^2$ = 17.2182, $p$ < 0.01

Test for subgroup differences (random effects): $\chi^2_1$ = 24.21, df = 1 ($p$ < 0.01)

#### FIGURE S3. ALL-CAUSE MORTALITY AMONG THOSE RECEIVING COVID-19 AND NON–COVID-19 VACCINES

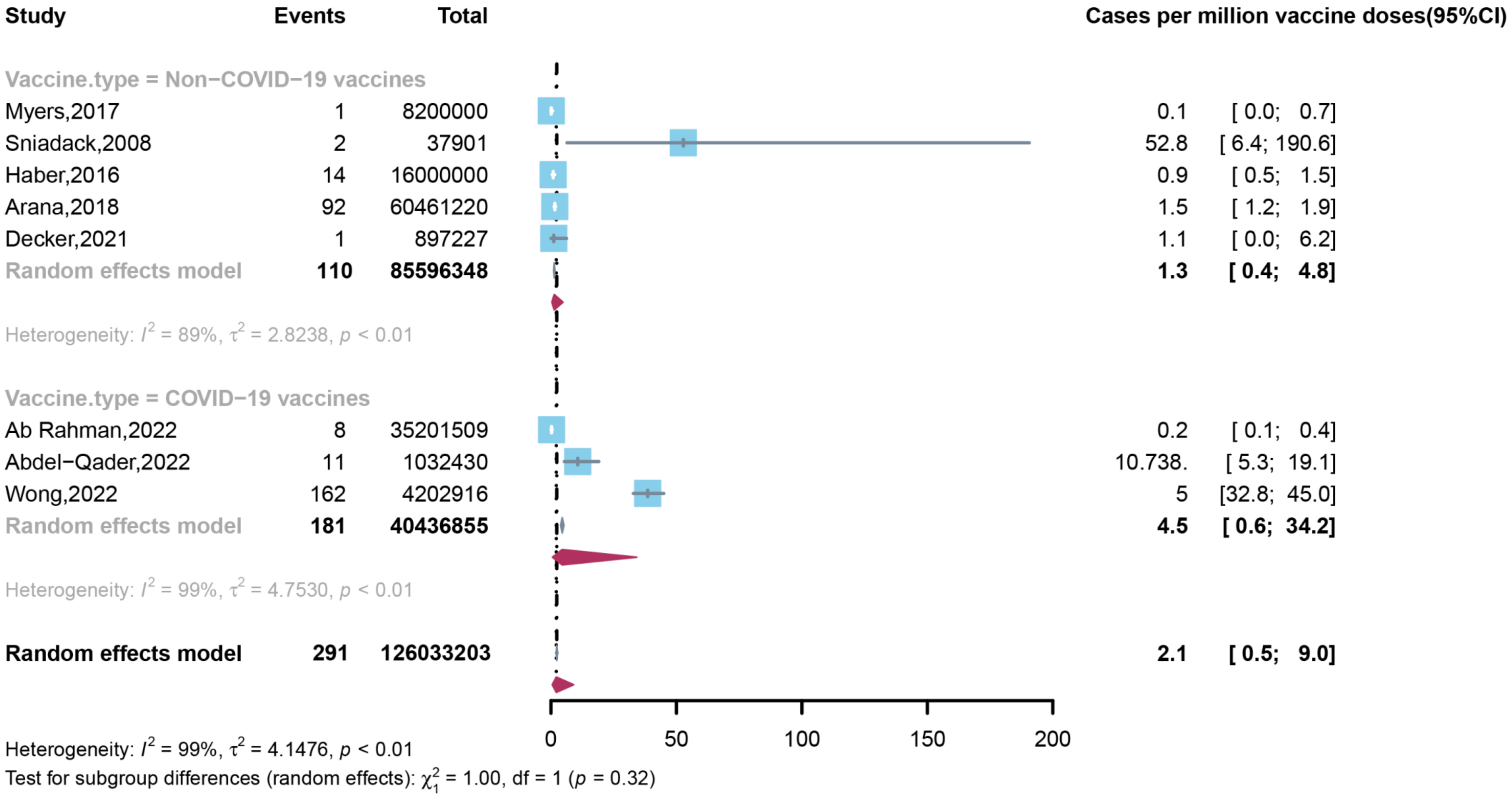

### FIGURE S4. INCIDENCE OF CARDIAC ARRHYTHMIAS AFTER COVID-19, HUMAN PAPILLOMAVIRUS, INFLUENZA, ACELLULAR PERTUSSIS, AND MIXED VACCINES

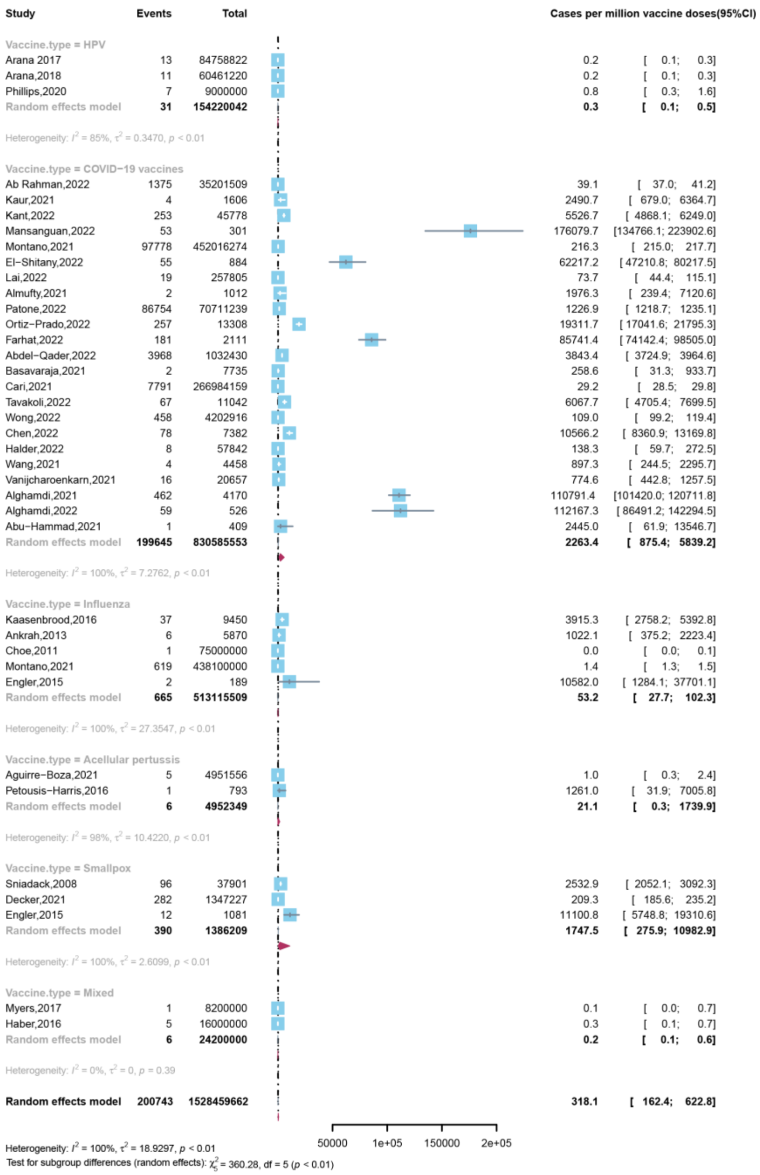

#### FIGURE S5. INCIDENCE OF CARDIAC ARRHYTHMIAS AFTER COVID-19 AND INFLUENZA VACCINES

| Study | Events | Total | Cases per million vaccine doses(95%CI) | |
|---|---|---|---|---|
| Vaccine.type = COVID−19 vaccines | | | | |
| Ab Rahman,2022 | 1375 | 35201509 | 39.1 | [ 37.0; 41.2] |
| Kaur,2021 | 4 | 1606 | 2490.7 | [ 679.0; 6364.7] |
| Kant,2022 | 253 | 45778 | 5526.7 | [ 4868.1; 6249.0] |
| Mansanguan,2022 | 53 | 301 | 176079.7 | [134766.1; 223902.6] |
| Montano,2021 | 97778 | 452016274 | 216.3 | [ 215.0; 217.7] |
| El−Shitany,2022 | 55 | 884 | 62217.2 | [ 47210.8; 80217.5] |
| Lai,2022 | 19 | 257805 | 73.7 | [ 44.4; 115.1] |
| Almufty,2021 | 2 | 1012 | 1976.3 | [ 239.4; 7120.6] |
| Patone,2022 | 86754 | 70711239 | 1226.9 | [ 1218.7; 1235.1] |
| Ortiz−Prado,2022 | 257 | 13308 | 19311.7 | [ 17041.6; 21795.3] |
| Farhat,2022 | 181 | 2111 | 85741.4 | [ 74142.4; 98505.0] |
| Abdel−Qader,2022 | 3968 | 1032430 | 3843.4 | [ 3724.9; 3964.6] |
| Basavaraja,2021 | 2 | 7735 | 258.6 | [ 31.3; 933.7] |
| Cari,2021 | 7791 | 266984159 | 29.2 | [ 28.5; 29.8] |
| Tavakoli,2022 | 67 | 11042 | 6067.7 | [ 4705.4; 7699.5] |
| Wong,2022 | 458 | 4202916 | 109.0 | [ 99.2; 119.4] |
| Chen,2022 | 78 | 7382 | 10566.2 | [ 8360.9; 13169.8] |
| Halder,2022 | 8 | 57842 | 138.3 | [ 59.7; 272.5] |
| Wang,2021 | 4 | 4458 | 897.3 | [ 244.5; 2295.7] |
| Vanijcharoenkarn,2021 | 16 | 20657 | 774.6 | [ 442.8; 1257.5] |
| Alghamdi,2021 | 462 | 4170 | 110791.4 | [101420.0; 120711.8] |
| Alghamdi,2022 | 59 | 526 | 112167.3 | [ 86491.2; 142294.5] |
| Abu−Hammad,2021 | 1 | 409 | 2445.0 | [ 61.9; 13546.7] |
| **Random effects model** | **199645** | **830585553** | **2263.4** | **[ 875.4; 5839.2]** |
| Heterogeneity: $I^2 = 100\%$, $\tau^2 = 7.2762$, $p < 0.01$ | | | | |
| Vaccine.type = Influenza | | | | |
| Kaasenbrood,2016 | 37 | 9450 | 3915.3 | [ 2758.2; 5392.8] |
| Ankrah,2013 | 6 | 5870 | 1022.1 | [ 375.2; 2223.4] |
| Choe,2011 | 1 | 75000000 | 0.0 | [ 0.0; 0.1] |
| Montano,2021 | 619 | 438100000 | 1.4 | [ 1.3; 1.5] |
| Engler,2015 | 2 | 189 | 10582.0 | [ 1284.1; 37701.1] |
| **Random effects model** | **665** | **513115509** | **53.2** | **[ 27.7; 102.3]** |
| Heterogeneity: $I^2 = 100\%$, $\tau^2 = 27.3547$, $p < 0.01$ | | | | |
| **Random effects model** | **200310** | **1343701062** | **1167.3** | **[ 653.4; 2084.7]** |

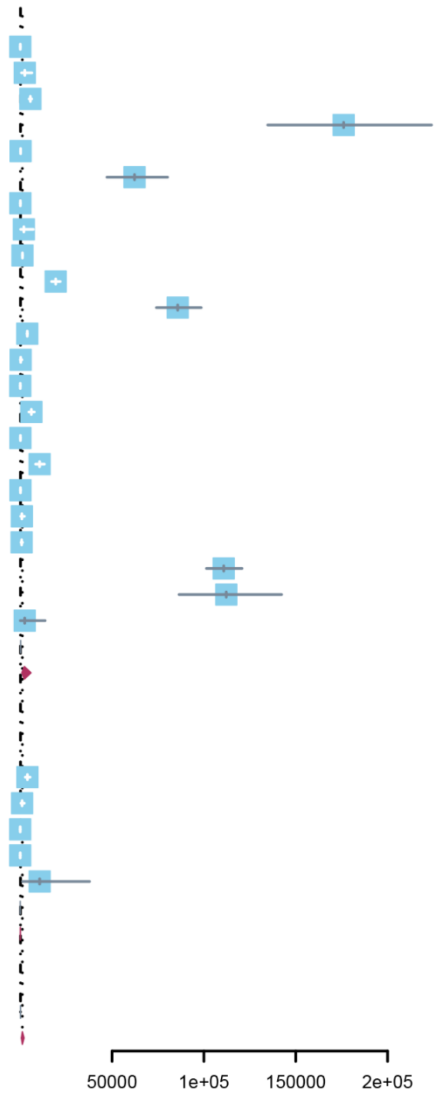

Heterogeneity: $I^2 = 100\%$, $\tau^2 = 12.5494$, $p < 0.01$
Test for subgroup differences (random effects): $\chi^2_1 = 40.62$, df = 1 ($p < 0.01$)

#### FIGURE S6. INCIDENCE OF CARDIAC ARRHYTHMIAS AFTER COVID-19 AND HUMAN PAPILLOMAVIRUS VACCINES

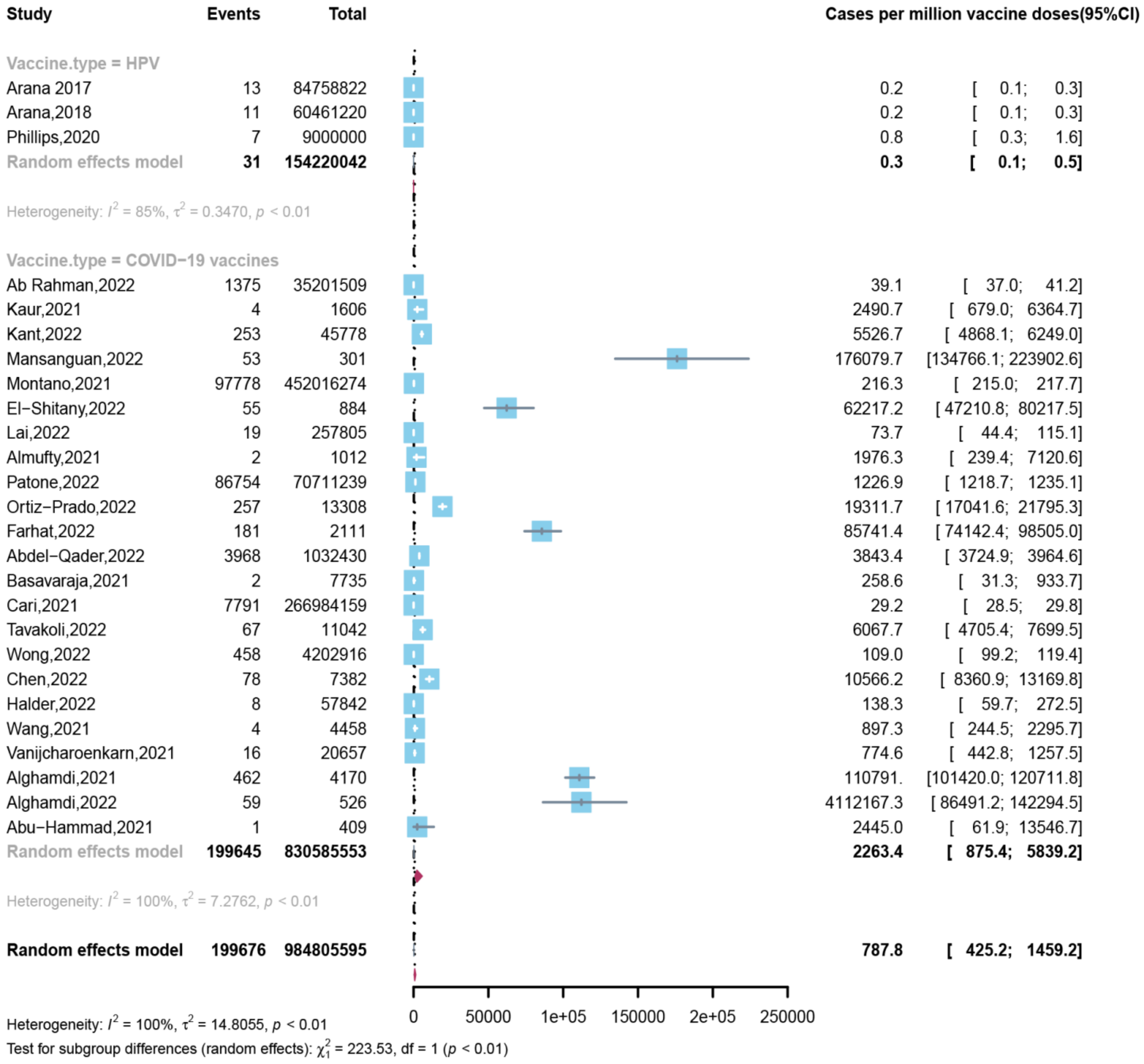

#### FIGURE S7. INCIDENCE OF CARDIAC ARRHYTHMIAS AFTER COVID-19 AND ACELLULAR PERTUSSIS VACCINES

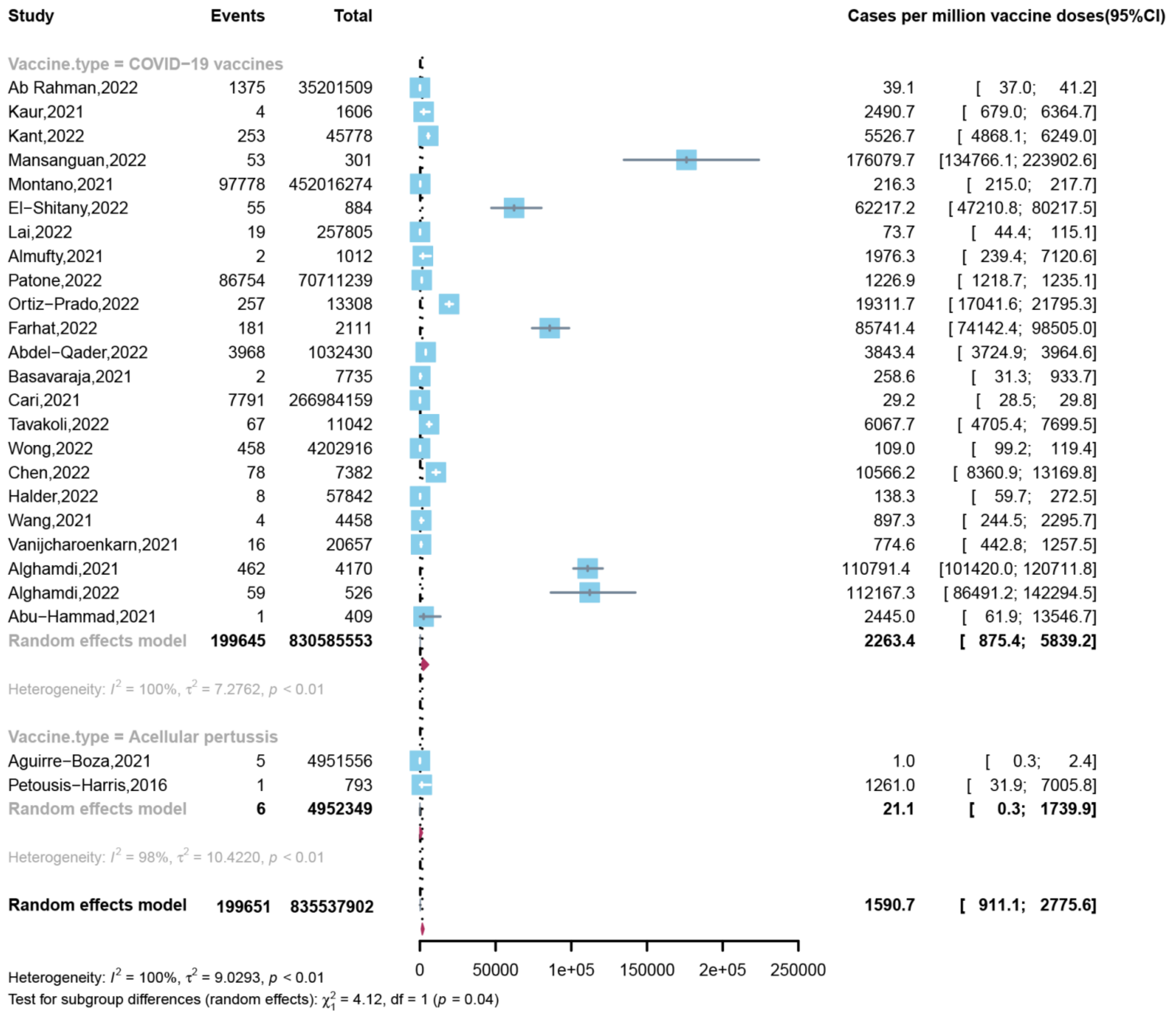

#### FIGURE S8. INCIDENCE OF CARDIAC ARRHYTHMIAS AFTER COVID-19 AND SMALLPOX VACCINES

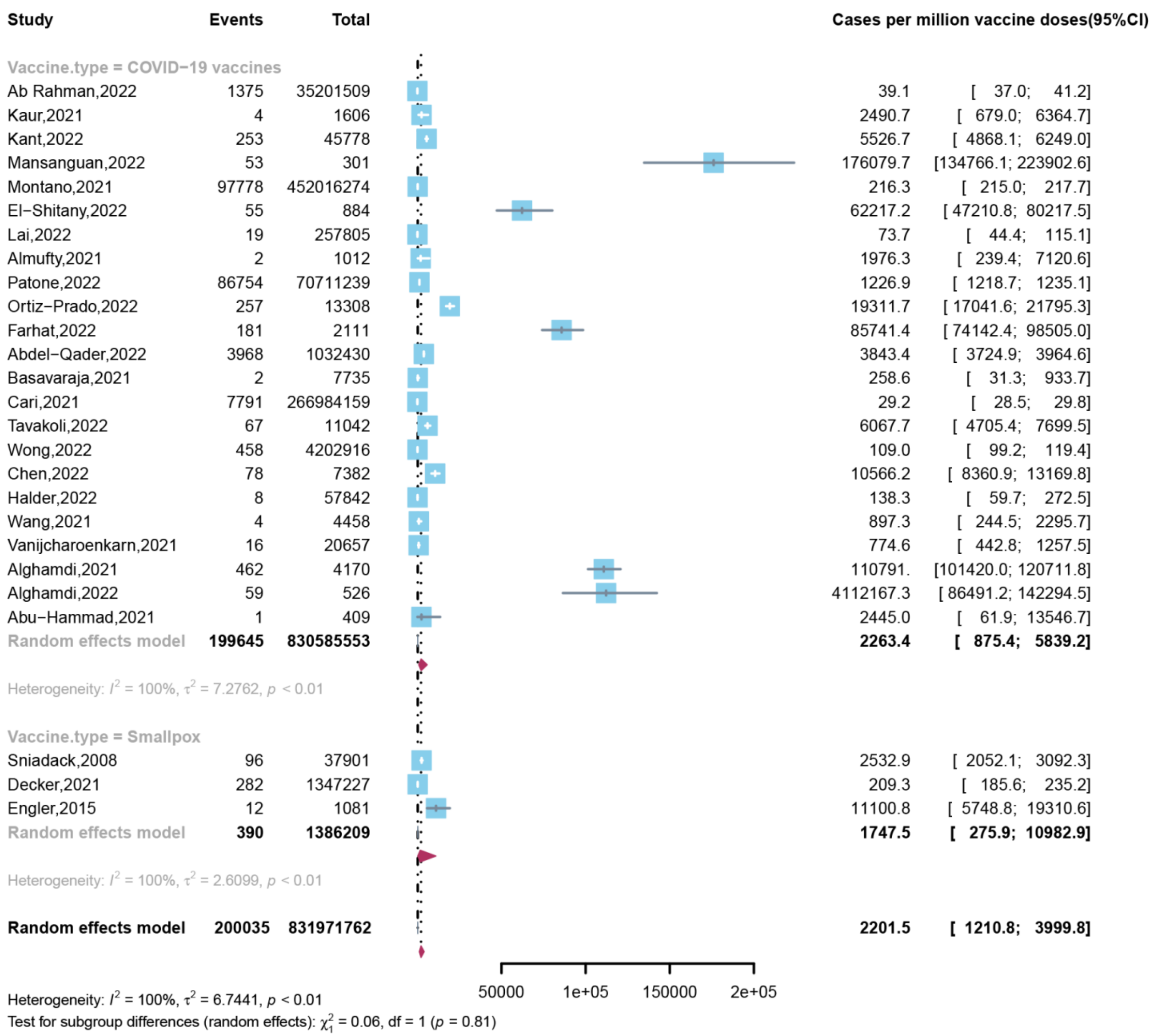

**FIGURE S9. INCIDENCE OF CARDIAC ARRHYTHMIAS AMONG THOSE RECEIVING mRNA COVID-19 AND NON–mRNA COVID-19 VACCINES**

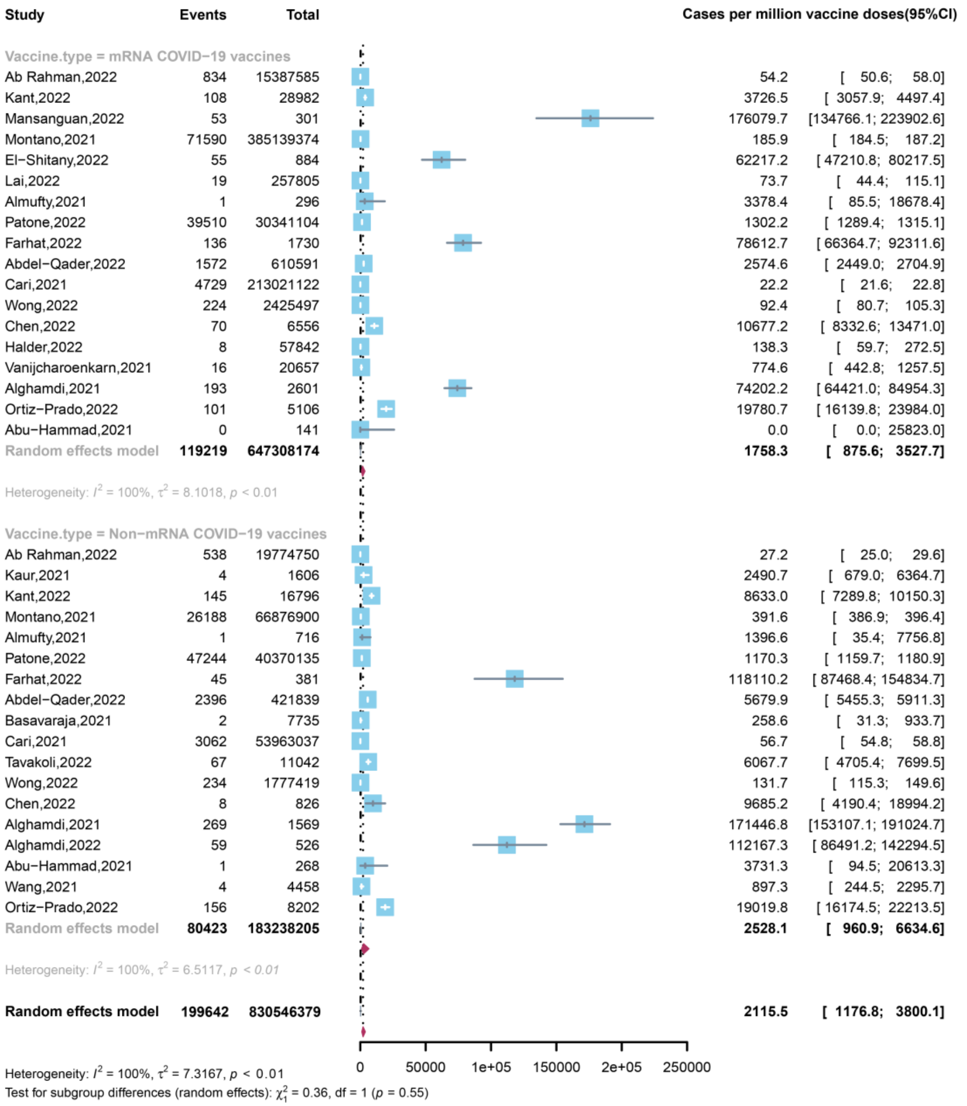

For the Ab Rahmam 2022 study, the total number of COVID-19 vaccines presented is lower than the reported number because some counts did not specify the type of COVID vaccine taken and therefore could not be included.

#### FIGURE S10. INCIDENCE OF CARDIAC ARRHYTHMIAS AMONG THOSE RECEIVING THEIR FIRST, SECOND, AND THIRD DOSE OF COVID-19 VACCINE

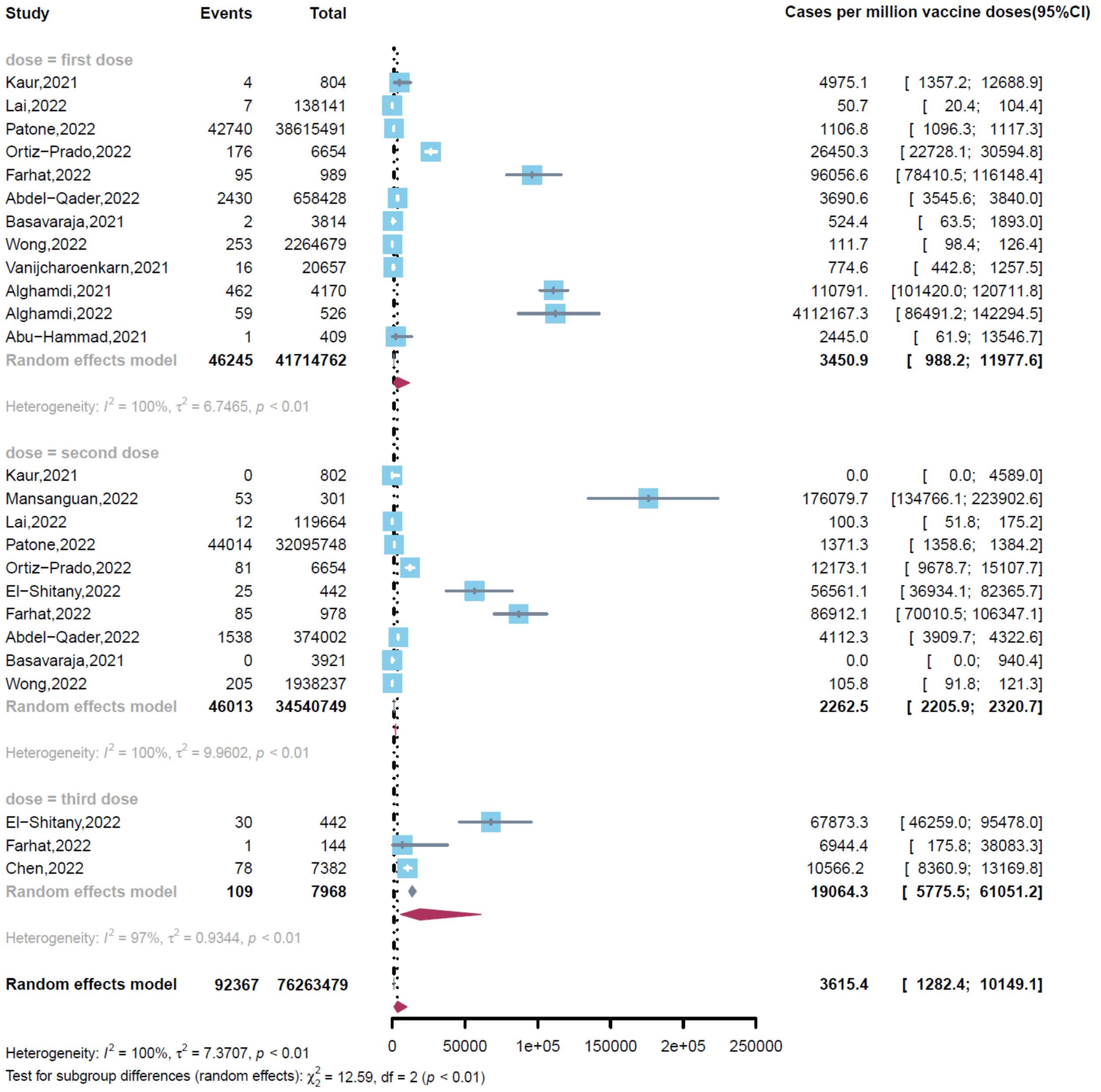

#### FIGURE S11. INCIDENCE OF CARDIAC ARRHYTHMIAS AFTER VACCINATION AMONG ADULTS AND CHILDREN

| Study | Events | Total | Cases per million vaccine doses | (95%CI) |
|---|---|---|---|---|
| Age = Adult | | | | |
| Kaasenbrood,2016 | 37 | 9450 | 3915.3 | [ 2758.2; 5392.8] |
| Sniadack,2008 | 89 | 37901 | 2348.2 | [ 1886.2; 2888.9] |
| Haber,2016 | 5 | 16000000 | 0.3 | [ 0.1; 0.7] |
| Ankrah,2013 | 6 | 5870 | 1022.1 | [ 375.2; 2223.4] |
| Kaur,2021 | 4 | 1606 | 2490.7 | [ 679.0; 6364.7] |
| Montano,2021 | 98397 | 890116274 | 110.5 | [ 109.9; 111.2] |
| Almufty,2021 | 2 | 1012 | 1976.3 | [ 239.4; 7120.6] |
| Abdel−Qader,2022 | 3968 | 1032430 | 3843.4 | [ 3724.9; 3964.6] |
| Cari,2021 | 7791 | 266984159 | 29.2 | [ 28.5; 29.8] |
| Basavaraja,2021 | 2 | 7735 | 258.6 | [ 31.3; 933.7] |
| Engler,2015 | 14 | 1270 | 11023.6 | [ 6039.5; 18426.7] |
| Wong,2022 | 458 | 4202916 | 109.0 | [ 99.2; 119.4] |
| **Random effects model** | **110773** | **1178400623** | **412.8** | **[ 269.7; 631.7]** |
| Heterogeneity: $I^2$ = 100%, $\tau^2$ = 7.6186, $p$ < 0.01 | | | | |
| Age = Children | | | | |
| Aguirre−Boza,2021 | 5 | 4951556 | 1.0 | [ 0.3; 2.4] |
| Mansanguan,2022 | 53 | 301 | 176079.7 | [134766.1; 223902.6] |
| Lai,2022 | 19 | 257805 | 73.7 | [ 44.4; 115.1] |
| Tavakoli,2022 | 18 | 11042 | 1630.1 | [ 966.4; 2575.1] |
| **Random effects model** | **95** | **5220704** | **390.7** | **[ 9.2; 16394.1]** |
| Heterogeneity: $I^2$ = 100%, $\tau^2$ = 20.1679, $p$ < 0.01 | | | | |
| **Random effects model** | **110868** | **1183621327** | **408.0** | **[ 185.6; 896.6]** |

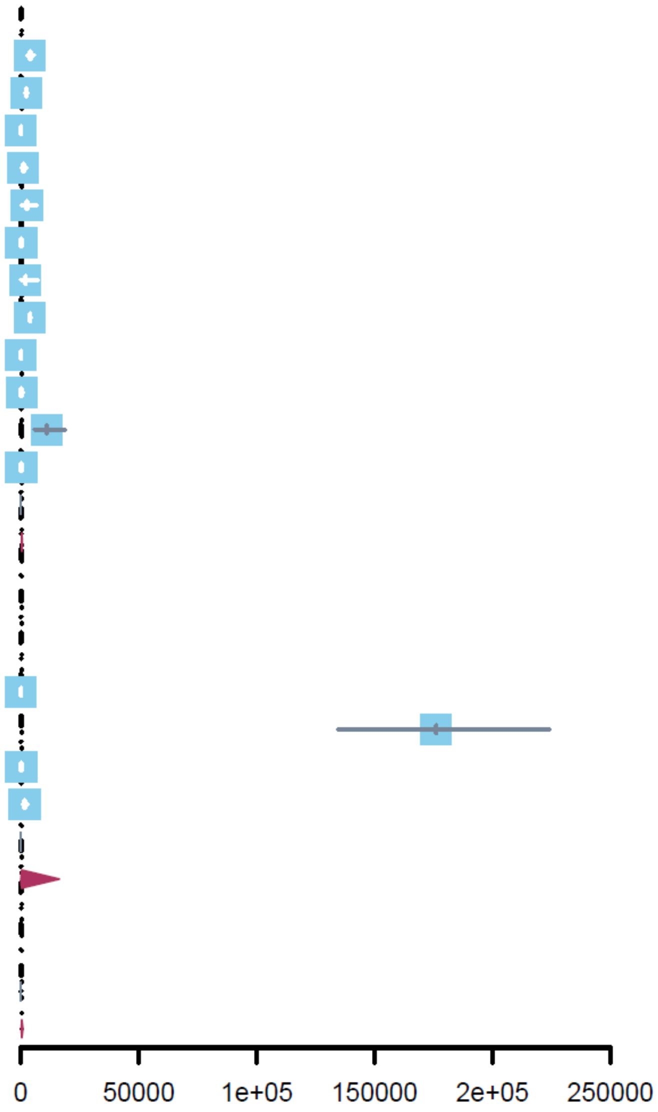

Heterogeneity: $I^2$ = 100%, $\tau^2$ = 10.7702, $p$ < 0.01
Test for subgroup differences (random effects): $\chi^2_1$ = 0.00, df = 1 ($p$ = 0.98)
